# Antidepressant efficacy of administering repetitive transcranial magnetic stimulation (rTMS) concurrently with psychological tasks or interventions: a scoping review and meta-analysis

**DOI:** 10.1101/2024.08.28.24312728

**Authors:** Cristian G. Giron, Alvin H.P. Tang, Minxia Jin, Georg S. Kranz

## Abstract

Current approaches to optimize the efficacy of repetitive transcranial magnetic stimulation (rTMS) for depressive symptoms focus on personalizing targets and parameters. But what should occur during these three-to-forty-minute sessions remains under-investigated. Specific concerns include evidence suggesting brain state modulates the brain’s response to stimulation, and the potential to boost antidepressant efficacy by administering rTMS concurrently with psychological methods. Thus, conducted a scoping review and meta-analysis, per PRISMA-ScR guidelines, to pool studies that administered rTMS during psychological tasks or interventions. PubMed and Web of Science databases were searched from inception to 10 July 2024. Inclusion criteria: neuropsychiatric patients underwent rTMS; studies assessed depressive symptom severity; psychological tasks or interventions were administered during rTMS, or intentionally did not include a wash-out period. Of 8442 hits, 20 studies combined rTMS with aerobic exercise, bright light therapy, cognitive training or reactivation, psychotherapy, sleep deprivation, or a psychophysical task. Meta-analyses with random effects models pooled the efficacy of these combinations, based on change scores on depressive severity scales. The effect size was large and therapeutic for uncontrolled pretest-posttest comparisons (17 studies, 20 datasets, g=-1.91, SE=0.45, 95%CI= −2.80 to −1.03, p<0.01); medium when studies compared active combinations with sham rTMS plus active psychological methods (8 studies, g=-0.55, SE=0.14, 95%CI= −0.82 to −0.28, p<0.01); and non-significant when active combinations were compared with active rTMS plus sham psychological methods (4 studies, p= 0.96). These findings suggest that the antidepressant efficacy of combining rTMS with psychological methods is promising, but not an improvement over rTMS alone.

## Introduction

Repetitive transcranial magnetic stimulation (rTMS) is a multi-tool for clinical practice and psychological research, with recommended applications across neuropsychiatric conditions As an antidepressant, the efficacy of rTMS is supported by meta-analyses for major depressive disorder (MDD; (2)), and across neuropsychiatric conditions when applied over the left dorsolateral prefrontal cortex (3). Strategies to improve the efficacy and consistency of rTMS protocols focus on personalizing stimulation targets or parameters that maximize the dose of stimulation (4, 5). But what should occur during treatment sessions has received much less attention. What patients do during sessions is typically not reported, and advice is not provided by expert guidelines (1) or cannot be established due to a lack of evidence (6). When details are reported, patients are instructed to relax, e.g., “During 10 Hz rTMS or iTBS sessions, participants were instructed to keep their eyes open and relax” (7). But expecting patients with depressive syndromes to consistently follow these instructions is problematic: internalizing disorders, such as depressive disorders, are characterized by emotional and cognitive dyscontrol linked to abnormal brain activity and connectivity (8, 9). Since distinct behaviors and thoughts correspond to different brain states (i.e., activity and connectivity), and as different brain states may underlie receptivity to stimulation, it has been suggested that patients’ actions and thoughts during stimulation contribute to the highly variable outcomes of rTMS (10–12). Such heterogeneity poses a significant challenge for antidepressant rTMS therapies. For instance, a meta-analysis of gold-standard trials applying rTMS to the left DLPFC found a large effect size but also substantial heterogeneity, indicated by a Higgin’s I^2^ of 86% (3). Further, inconsistent changes in brain hemodynamics are observed in both the targeted prefrontal region and remote areas during and after rTMS (13).

### Evidence for the ‘brain state’ hypothesis of rTMS

The potential for concurrent psychological tasks or interventions to alter the brain’s response to rTMS, potentially controlling for brain state, is of great interest to clinicians and researchers. In reviews focusing on the cognitive neuroscience literature, Silvanto and colleagues suggest that TMS does not broadly excite, inhibit, or induce “virtual lesions” of targeted brain regions (12, 14). Instead, neuronal pools within these regions respond distinctly to stimulation, depending on their activity during stimulation. Similar reasoning has driven experiments to explore whether behavior during rTMS modulates brain responses in healthy participants. Neuroimaging studies show that the after-effects of high-frequency and high-intensity rTMS over the left dorsal premotor cortex are excitatory when the left hand is actively gripping during stimulation, but inhibitory when the left hand is at rest (15). Similar effects were observed using continuous and intermittent theta burst stimulation protocols (16). In another study, Luber and colleagues (17) paired audio-visual stimuli with single pulses in a classical conditioning task. The presentation of conditioned stimuli, but not unconditioned stimuli, before TMS attenuated motor-evoked potentials. More recently, a study interleaving TMS-fMRI found varied blood-oxygenation changes in response to TMS depending on whether healthy participants were at rest or performing the n-back task (18). These neuroimaging studies with healthy participants support the hypothesis that brain state at the time of TMS modulates the brain response to stimulation. However, behavioral studies do not support this hypothesis (19–21). To wit, behavioral after-effects following high frequency rTMS of the left dorsolateral prefrontal cortex (DLPFC) are not modulated when healthy participants viewed positive compared to neutral affect-inducing images during stimulation (19), nor when stimulation is delivered during a working memory task compared to rTMS alone (20). However, these behavioral studies assessed after-effects with task performance or self-reported mood items that may not be sufficiently sensitive. Indeed, meta-analyses of studies with healthy participants report that rTMS alone has null or little effects on these behavioral measures (22, 23).

### Previous reviews or perspectives on brain states and rTMS

The evidence supports the hypothesis that rTMS after-effects can be influenced by participants’ thoughts or actions during stimulation. This possibility has attracted significant interest from researchers and clinicians, leading to several reviews and perspectives on the topic (10, 11, 24–28). However, these have been non-systematic narrative reviews or overviews (10, 11, 24, 26, 27), did not examine treatment efficacy (26), or did not consider on the timing of the combined interventions (24, 25, 27, 28). We determined a scoping review was necessary to charter this literature and estimate efficacy, as we expected the literature to be highly heterogeneous inherent of rTMS research (2, 3), compounded by the variability of psychological tasks, interventions, and their timing with rTMS. To our knowledge, this is the first systematic effort to chart and conduct a meta-analysis on this research question. We aim to ground these findings by comparing efficacy estimates with published meta-analyses on the efficacy of rTMS alone and conduct power analyses to guide future research.

### Objectives

This scoping review systematically charts and meta-analyzes literature on the antidepressant efficacy of rTMS administered during or immediately after psychological tasks and interventions – putatively, rTMS with brain state manipulated. We included studies that combined these methods to treat any neuropsychiatric condition and reported effects on depression severity. Findings are discussed to inform combination designs for clinical and experimental research. The specific research objectives are as follow:

1. To investigate whether the antidepressant effects of rTMS are modulated (i.e., identify synergistic or antagonistic effects) by combination with psychological tasks and interventions.
2. Charter the literature relevant to future clinical and research paradigms integrating clinical rTMS protocols with psychological methods

## Methods

This scoping review complies with the reporting guidelines of the Preferred Reporting Items for Systematic Reviews and Meta-Analyses extension for Scoping Reviews (PRISMA-ScR) (29). Its protocol was registered with the Open Science Framework on 25 June 2022 (https://osf.io/n8hw3).

### Eligibility criteria

The PICO model summarizing inclusion criteria is as follows:

- **Patient:** Human neuropsychiatric patients with depressive symptoms. Treated patients do not need to have a depressive disorder as a primary diagnosis. For example, if treated patients are diagnosed with post-traumatic stress disorder or a pain disorder, the study can be included if depression severity was assessed.
- **Intervention:** Non-pharmacological intervention overlaps with sessions of rTMS. Chronotherapies can be included if wash-out periods are not used (e.g., patients are sleep deprived during days where rTMS is administered). Combined neuromodulation or open- and closed-loop designs are excluded, as this literature is reviewed elsewhere. Studies are also excluded if only pharmacotherapy is combined with rTMS.
- **Comparison:** No restriction.
- **Outcome:** Study reports depression severity such as with the Hamilton Depression Rating Scale, visual analogue scales, or assessments by trained clinicians

Excluded criteria were: 1) rTMS was administered alone; 2) timing of treatments were not concurrent with rTMS; 3) combined rTMS with pharmacotherapy only; 4) combined rTMS with other neuromodulatory devices (e.g., transcranial electric stimulation), or utilized an open- or closed-loop design without a psychological task or intervention.

### Information sources and search strategy

The search was performed using broad terms for rTMS and depression in the PubMed database until 5 May 2022 (YTC, HYCC, SWC, YYC) and updated until 10 July 2024 to also include Web of Science database (CGG, AHPT, MJ). Search queries used for each database are shown in **Supplementary Table S1**. Search results were exported into EndNote 20 to facilitate screening. This was done independently by separate reviewer teams (YTC, HYCC and SWC, YYC), then updated by a separate team (CGG and AHPT, MJ). Reference list of relevant reviews and included studies were also screened. We anticipated screening would require multiple teams since the search queries and inclusion criteria were intentionally broad. Furthermore, reporting what patients do during rTMS sessions is unstandardized in this literature. Thus, many studies were expected to need careful full-text screening to confirm whether psychological tasks or interventions were combined with rTMS, and if so, whether they were concurrent.

### Study selection and data chartering

Reviewers first screened by title and abstract independently, then discussed and merged these screening results. Full-text retrieval and screening was then conducted by the same independent teams. Disagreements on study selection were resolved by discussion and consensus with GSK. No automation tools were used for full-text screening. A customized Excel spreadsheet was developed for data chartering by CGG (items are described in the next section). Once full-text screening was completed, data was extracted by CGG, YTC, HYCC, SWC, and YYC, then verified by MJ and AHPT. Any disagreements were resolved through discussion with GSK.

### Extracted data

The following items were extracted using a customized Excel spreadsheet: participant characteristics for each treatment group (diagnoses and diagnostic method; age; sex ratio); rTMS protocol (target site, pulse pattern and intensity, number of sessions); psychological task or intervention; timing of treatments; measurements of antidepressant outcome; reported results of statistical comparisons; estimated effect sizes for within-subjects and between-subjects comparisons for each study. If these data were visually presented but not numerically available, we used WebPlotDigitizer to extract numerical values (https://automeris.io/WebPlotDigitizer/, last accessed 23 June 2023). If needed efficacy data was not obtainable in the full-text or supplementary materials of a study, we contacted corresponding authors of a study via.

### Groups and Comparisons

To assess antidepressant efficacy, change scores were calculated for each group across treatment timepoint by subtracting baseline scores from endpoint scores on standardized clinical scales, such as the Hamilton Depression Rating Scale (HDRS). These change scores were then pooled with respect to comparison. Effect sizes were estimated for within-group comparisons (posttest minus pretest) and controlled comparisons (change scores of the active intervention minus control condition). We estimated separate meta-analytic effect sizes for the following comparisons, where PSYC refers to psychological task or intervention:

- Uncontrolled: [active rTMS + active PSYC] endpoint versus baseline scores;
- Controlled: [active rTMS + active PSYC] versus [active rTMS + sham PSYC];
- Controlled: [active rTMS + active PSYC] versus [sham rTMS + active PSYC];
- Controlled: [active rTMS + active PSYC] versus [sham rTMS + sham PSYC].

These comparisons are necessary tests for the hypothesis that combining treatments alters efficacy. For example, a combination of methods can be synergistic or antagonistic (i.e., a negative interaction effect). It is essential to consider controls for both rTMS and PSYC to make causal claims about altered efficacy following their combination.

For studies where statistical comparisons were not reported (e.g., case studies), post-treatment response or remission outcomes were extracted to estimate the antidepressant efficacy. Such studies were not included in any meta-analysis.

### Effect sizes based on change-scores of within-group and between-group comparisons

To compute effect sizes, we used custom scripts written in Python (version 3.11.5) with NumPy and panda libraries in a Jupyter Notebook environment. We computed standardized mean differences of change scores for individual study comparisons. Details of relevant algorithms (30, 31) and pseudocode is provided in **Supplementary Text 1**.

### Meta-analysis and tests of heterogeneity

Meta-analyses were conducted when at least three included studies conducted similar comparisons. We used the *metafor* package (32) in R version 4.3.3 (33) to conduct the meta-analyses and meta-regressions, to assess publication and sensitivity bias, and to conduct subgroup analyses described below. Random-effects models were used for all comparisons to account for predicted variability among study effects. The Restricted Maximum Likelihood (REML) method was used to estimate the between-study variance.

To assess heterogeneity, we used Cochran’s Q test and Higgin’s I^2^ statistic. A significant Q test indicates the presence of heterogeneity among study effects beyond sampling error. The I² statistic is used to estimate the proportion of total variation due to heterogeneity rather than sampling error: I² values below 30% indicate low heterogeneity, between 30-60% indicate moderate heterogeneity, values above 60% indicate substantial heterogeneity (30).

### Publication and sensitivity bias analyses

Publication bias was assessed using funnel plots and Egger’s regression intercept test in meta-analysis with ten or more studies due to concerns about insufficient power (34). Leave-one-out cross-validation sensitivity tests were conducted to identify whether any single study disproportionately influences meta-analytic results. To wit, we iteratively excluded one study at a time, re-ran the meta-analysis using *metafor* in R, and observed how the results changed with the exclusion of the given study. To assess the robustness of each meta-analytic estimate, we report: the range of Hedges’ g’s observed; highlight whether the direction of effect changes; and whether the meta-analytic estimate remains significant.

### Meta-regression

To assess potential moderators, we conducted meta-regressions for meta-analytic estimates consisting of ten or more studies (30). The assessed moderators were sex ratios (number of male patients divided by number of female patients in a sample), mean age of a sample, and whether the psychological task or intervention was a supported intervention for depressive symptom (e.g., psychotherapies) or not (e.g., psychophysical tasks).

### Secondary meta-analysis

Meta-analyses were repeated to include only studies targeting the left dorsolateral prefrontal cortex (left DLPFC) and recruited patients with depressive disorders. This would allow us to compare meta-analytic findings with a recent cross-diagnostic meta-analysis that synthesized the effects of active rTMS relative to sham rTMS (3). This would allow us to compare combination treatment efficacy with rTMS alone.

### Monte-Carlo simulation-based power analysis

To estimate the sample size needed to obtain sufficient power (β=0.80, given ⍺=0.05) to investigate the hypothesis that combining rTMS with PSYC is a superior treatment (compared to sham conditions), we used the results of representative studies in simulation-based power analysis. Methods are described in **Supplementary Text 4**.

## Results

### Selection and sources of evidence

8442 records were identified in PubMed, Web of Science, and the reference lists of included studies and relevant reviews. The summary flow diagram is shown in **Figure 1**, and the PRISMA-ScR checklist is provided in **Supplementary Table S2**. References and reasons for 183 excluded studies that underwent full-text screening are shown in **Supplementary Table S3**. Twenty studies were eligible for inclusion.

**Figure 1.**
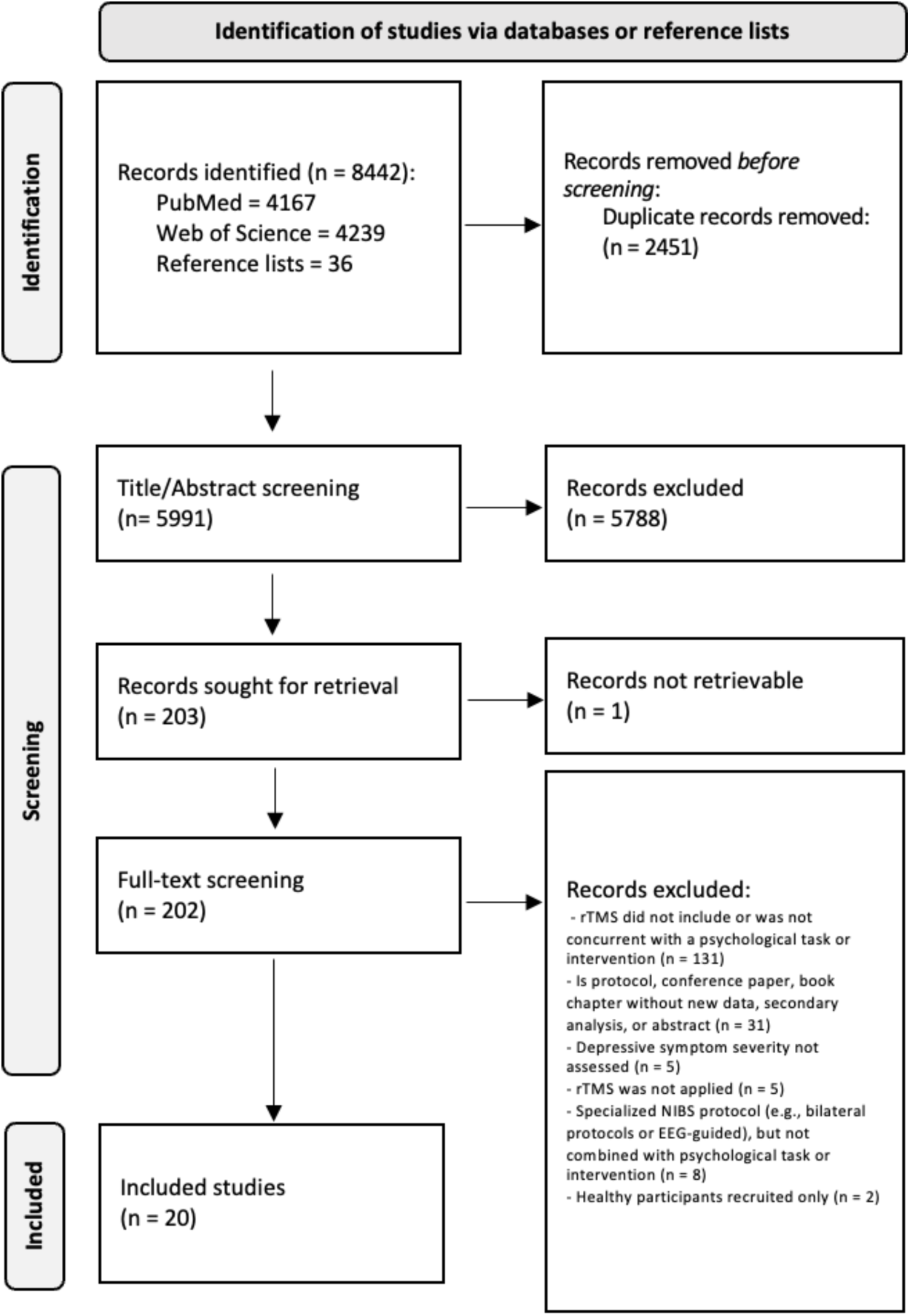
Summary flow-diagram of literature screening.

### Characteristics of included studies

The literature on modulating antidepressant outcomes of rTMS or psychological interventions by their combination comprised twenty studies (35–54), chartered in **Table 1**. Eleven studies conducted between-group comparisons (35, 40–45, 47, 49, 50, 52), six reported within-group comparison only (36, 38, 39, 46, 48, 51), and three were a case study or series (37, 53, 54)

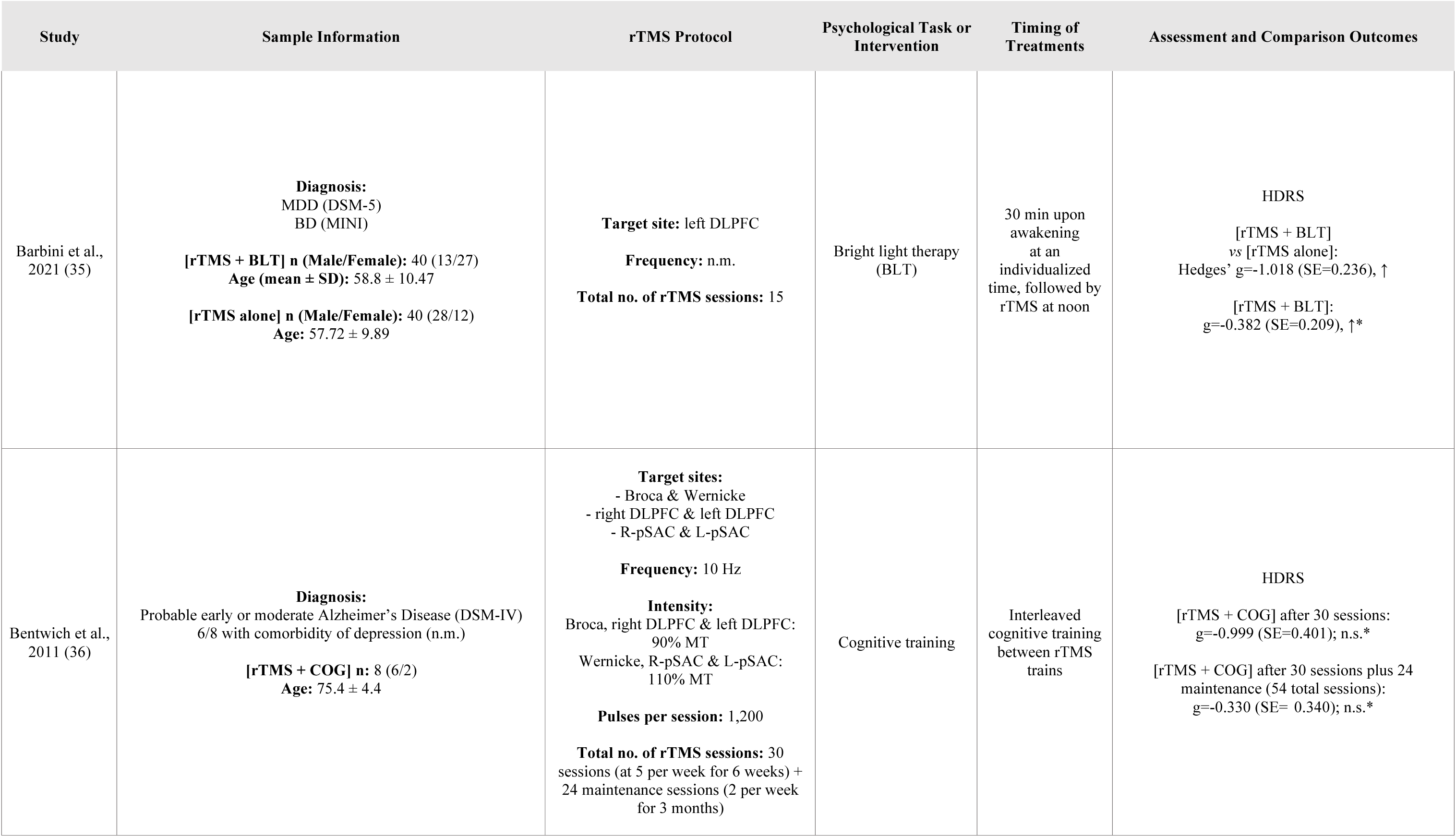
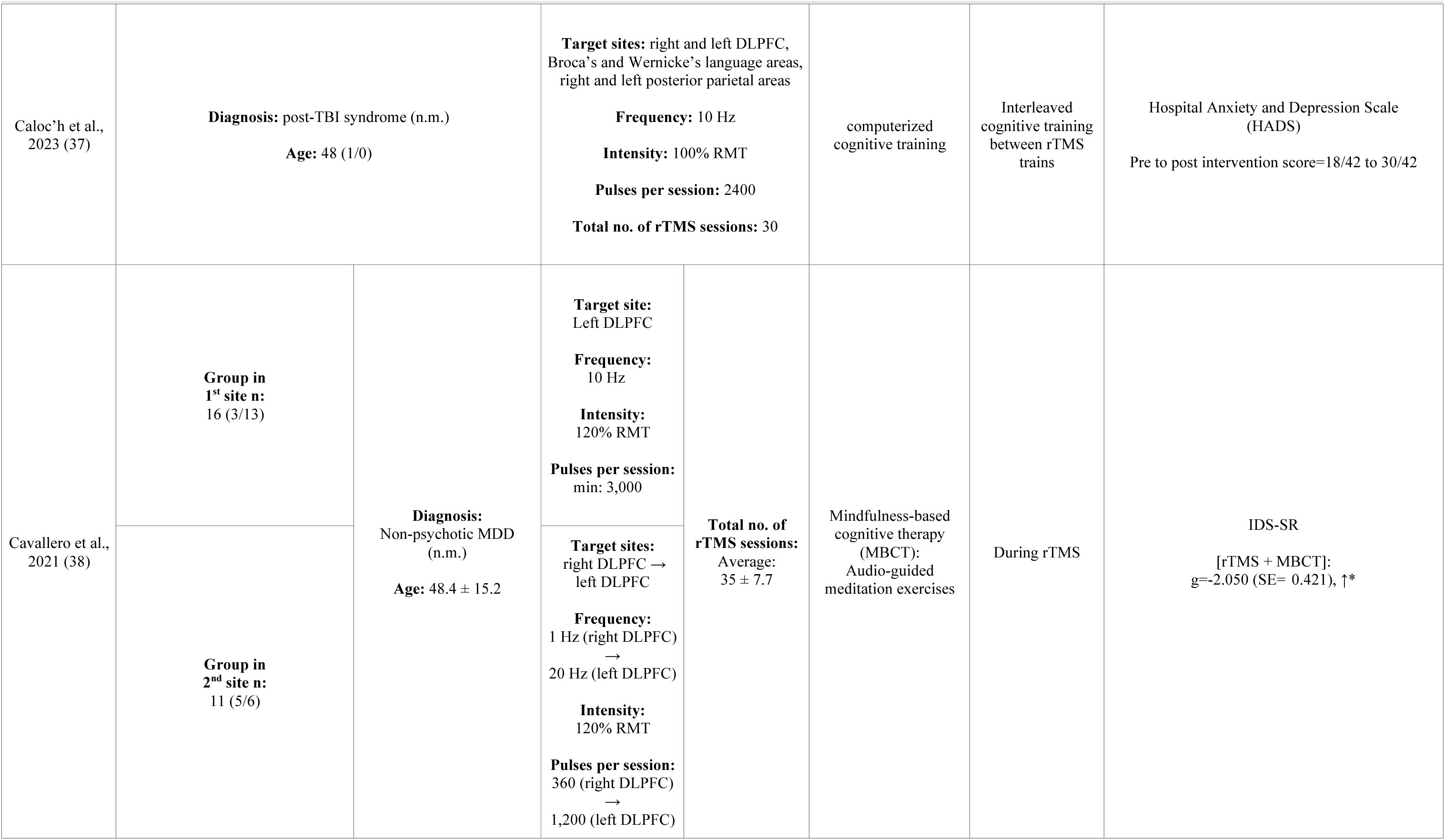
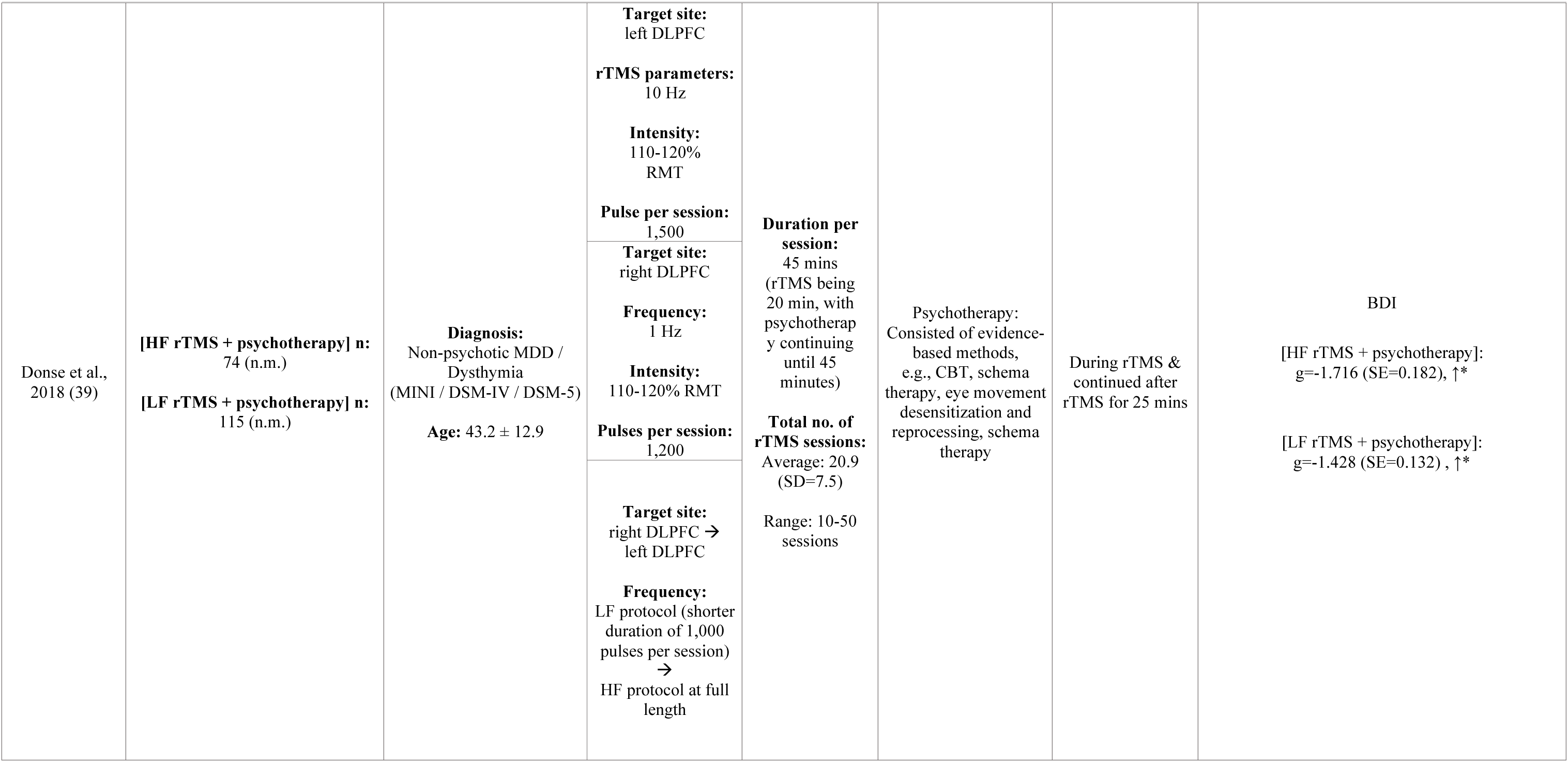
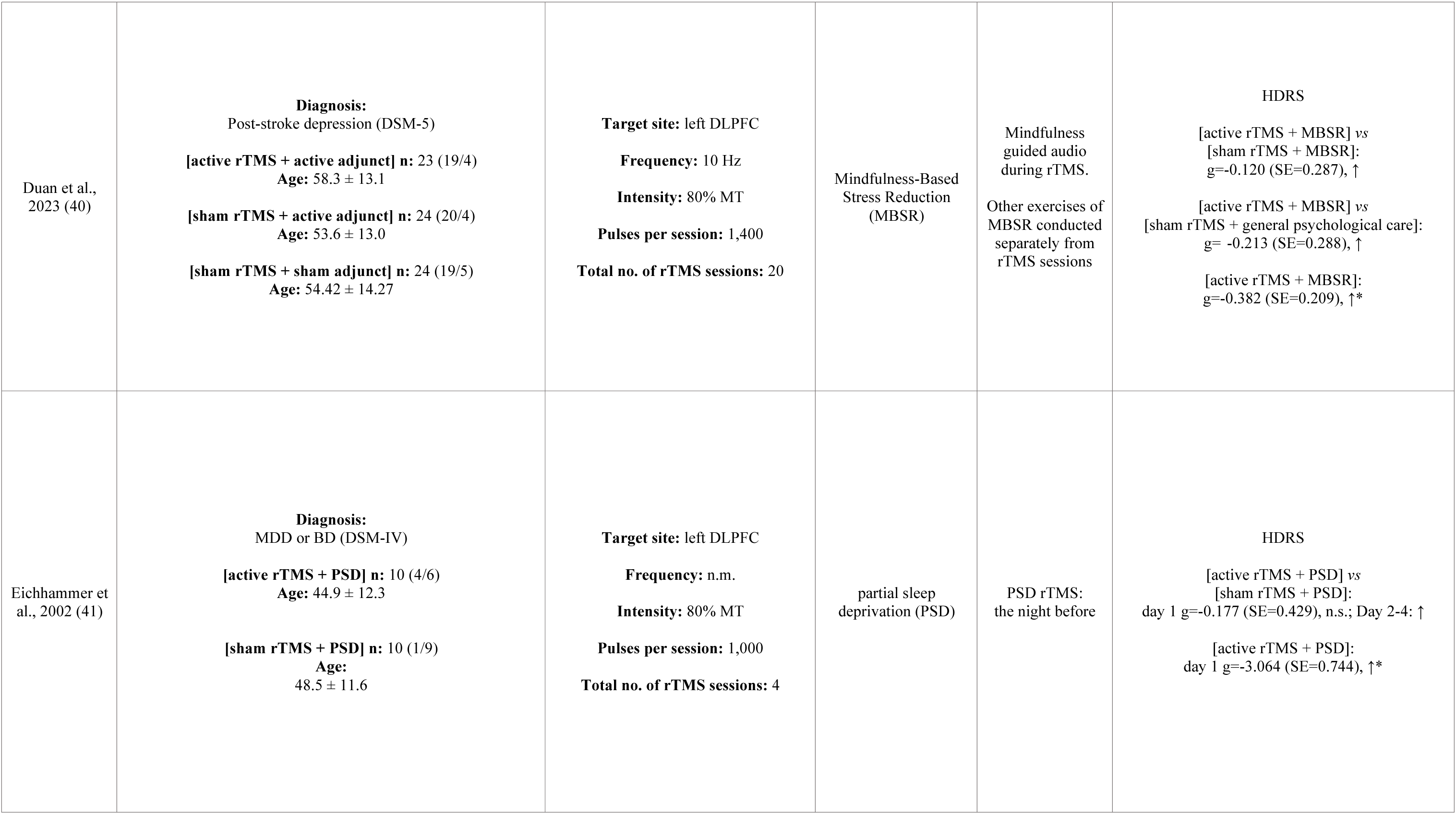
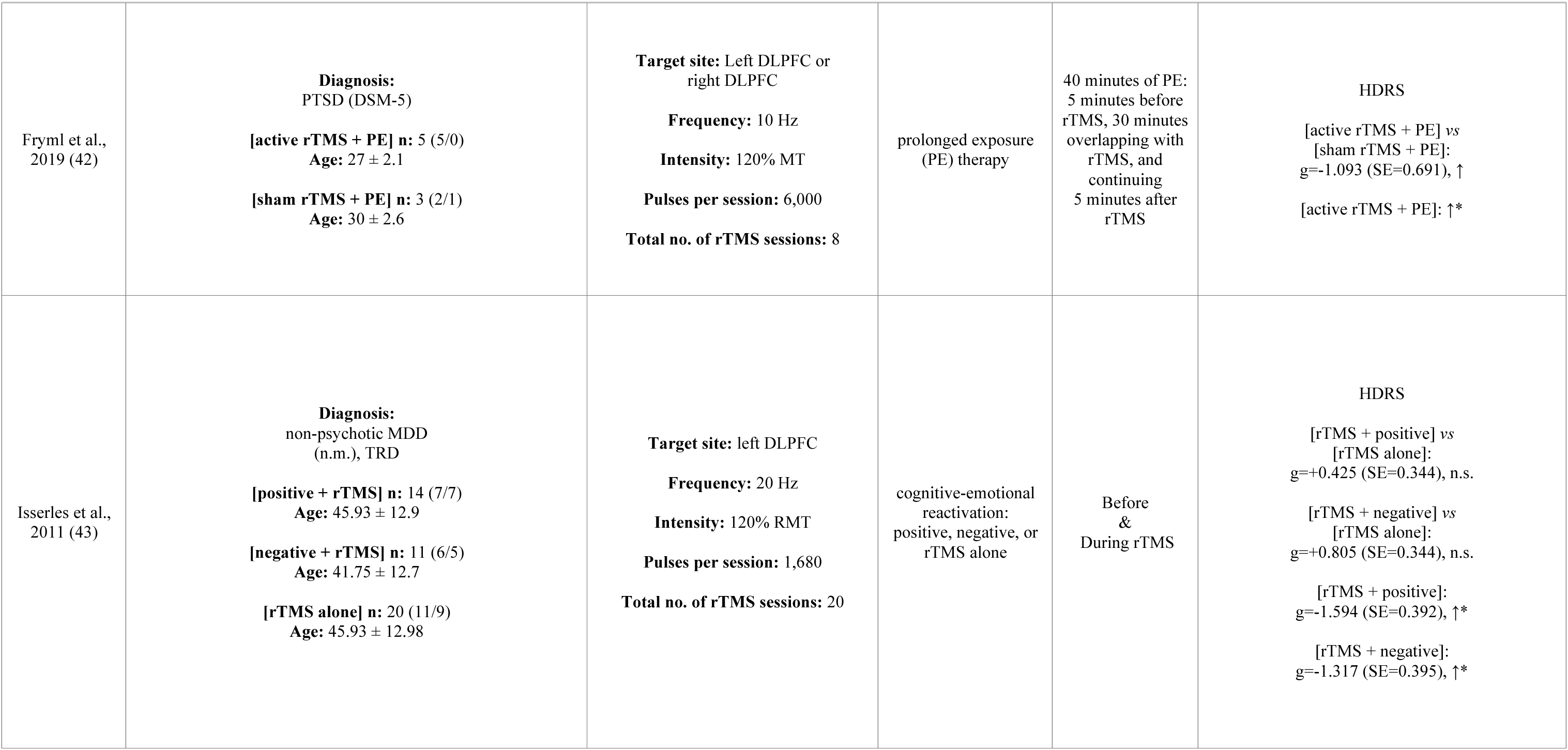
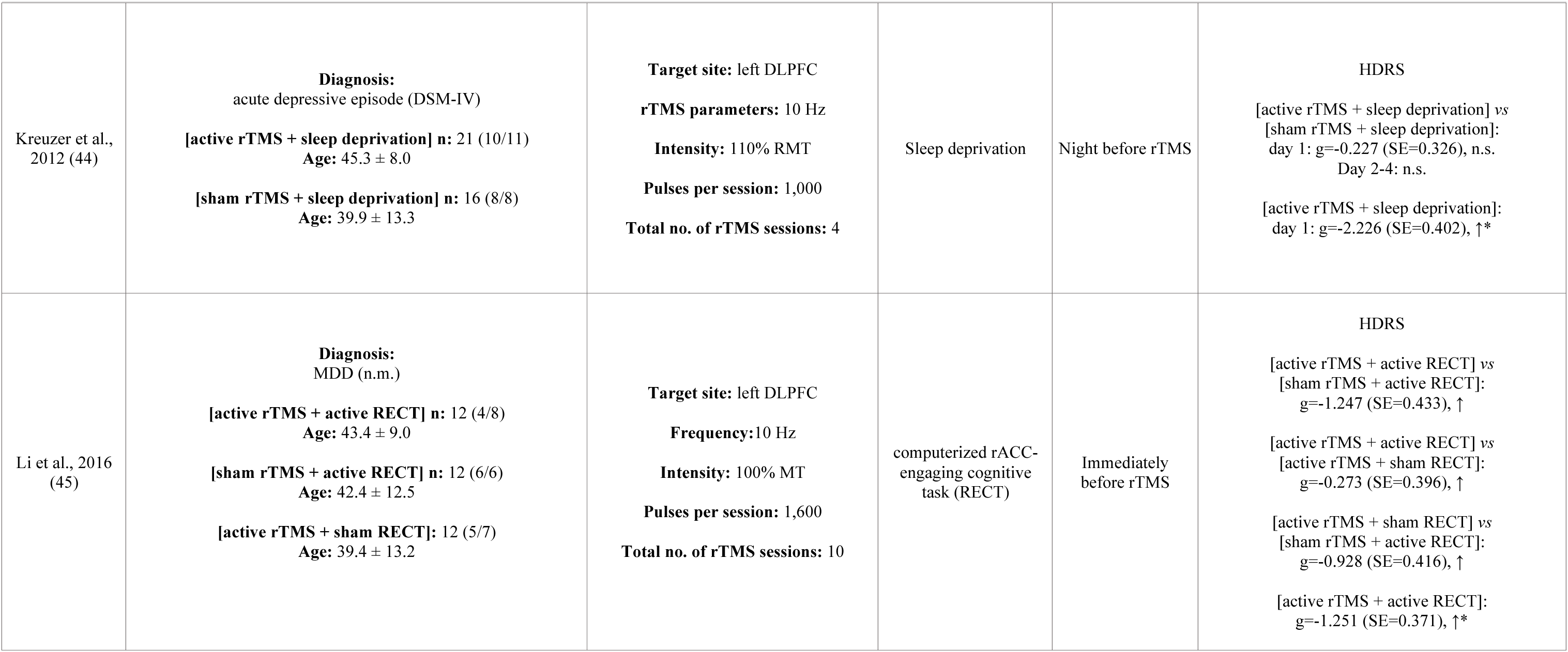
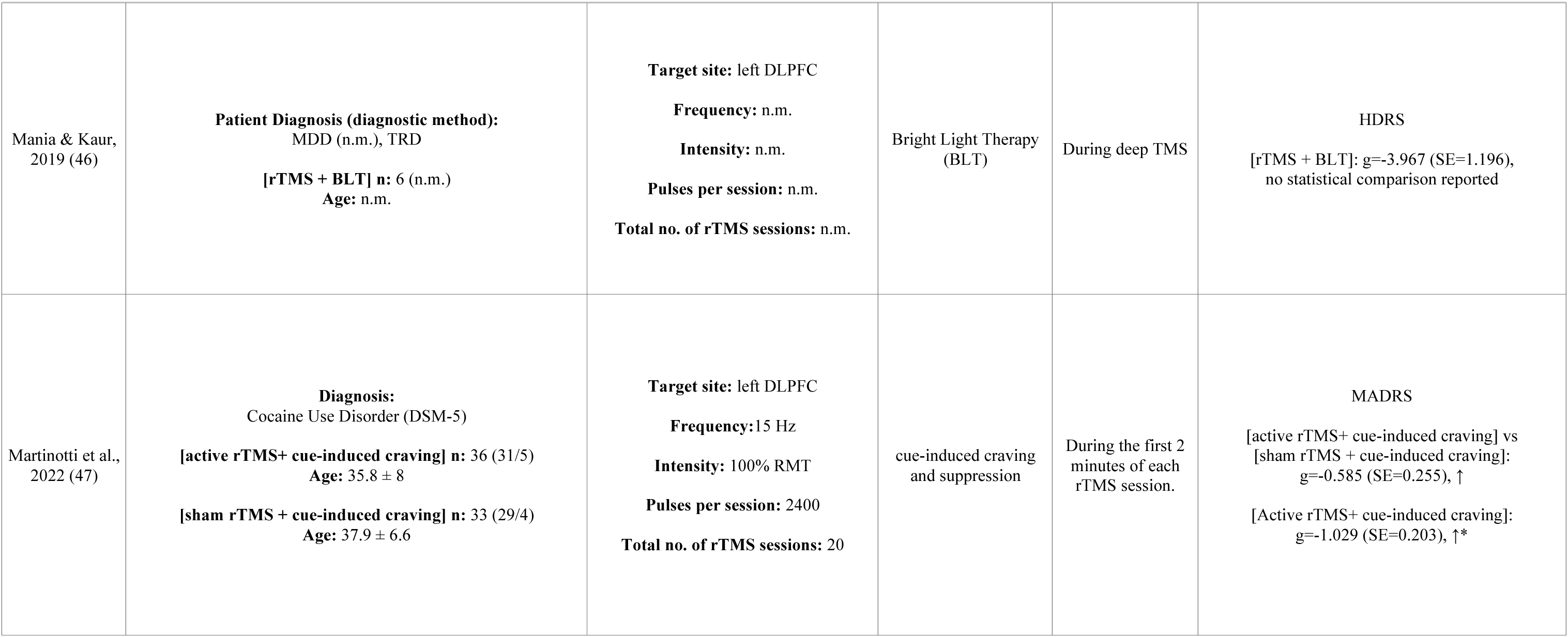
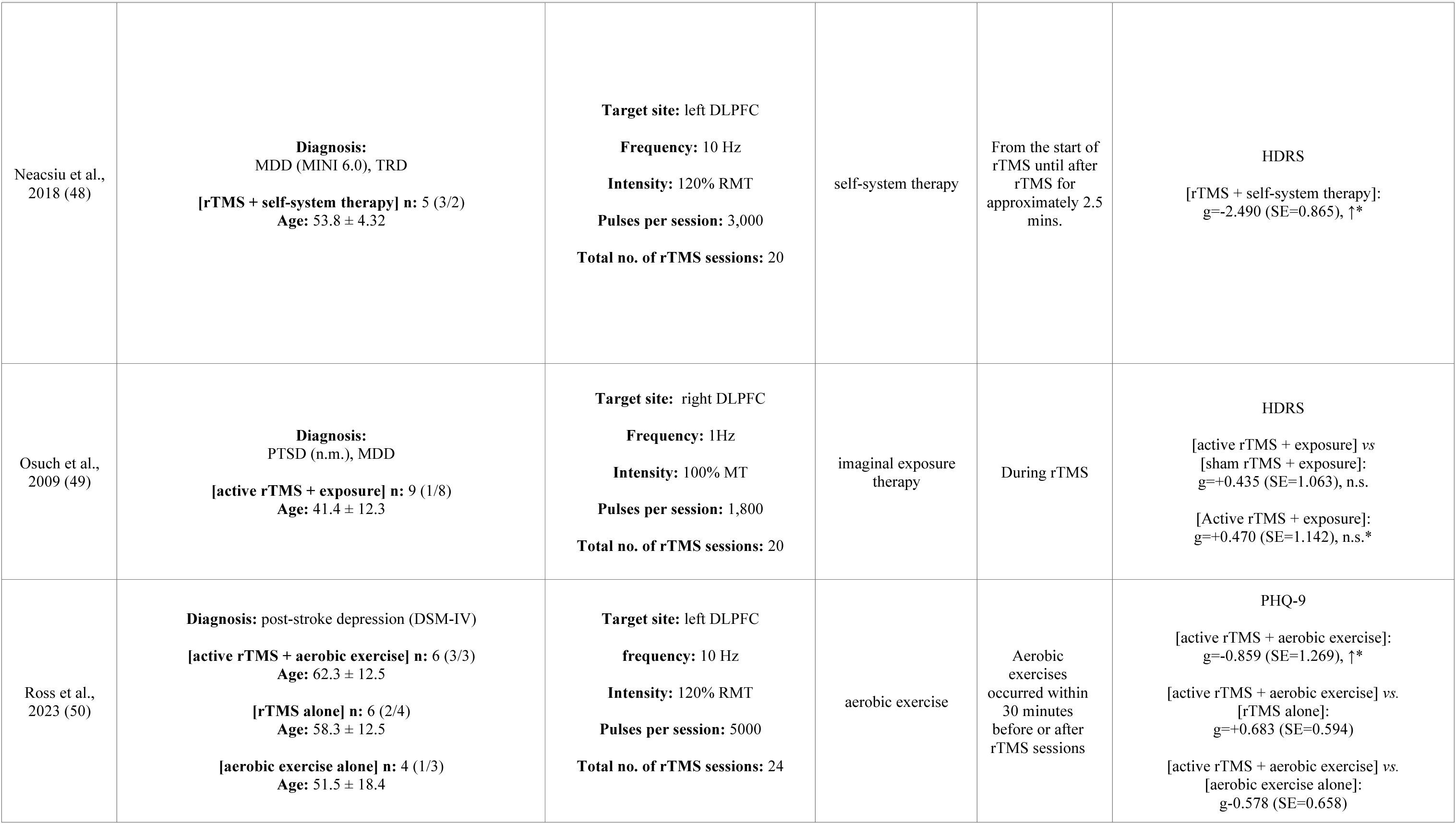
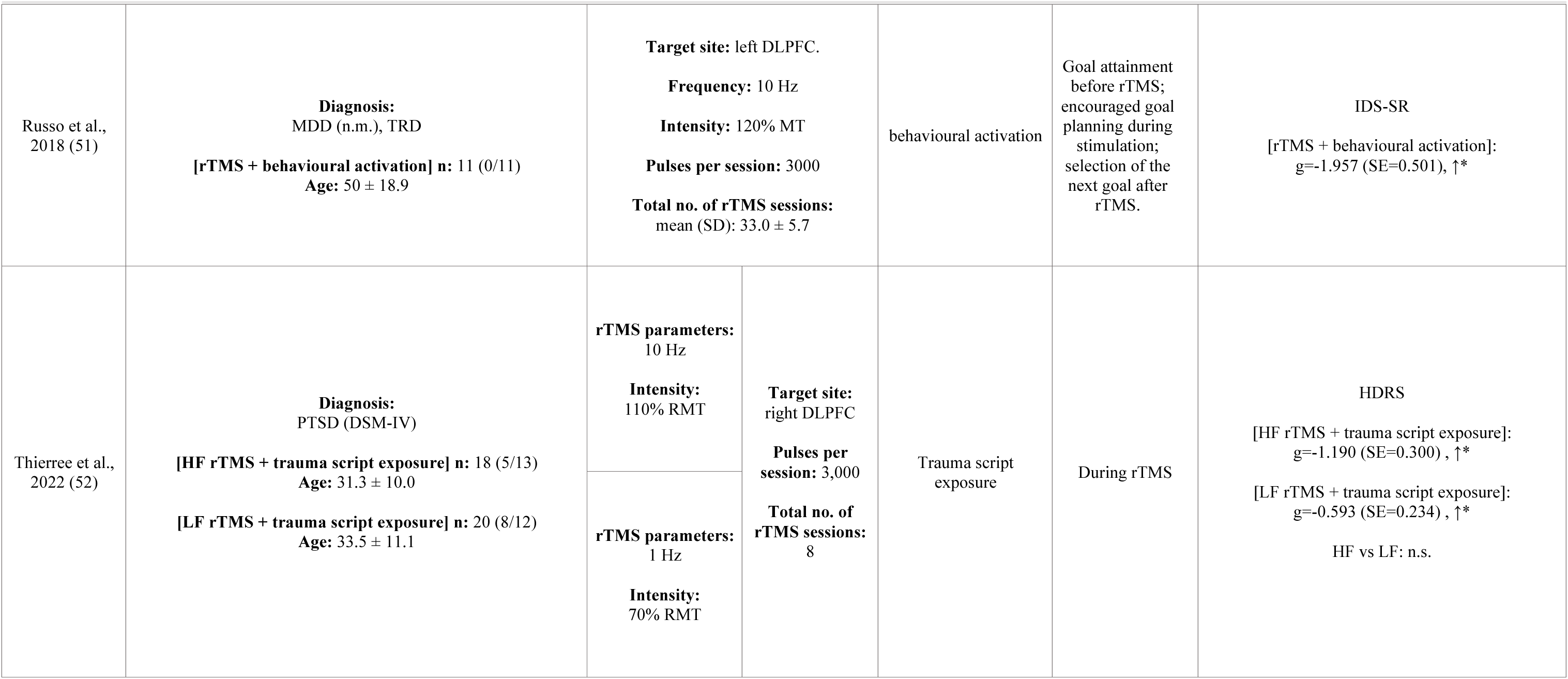
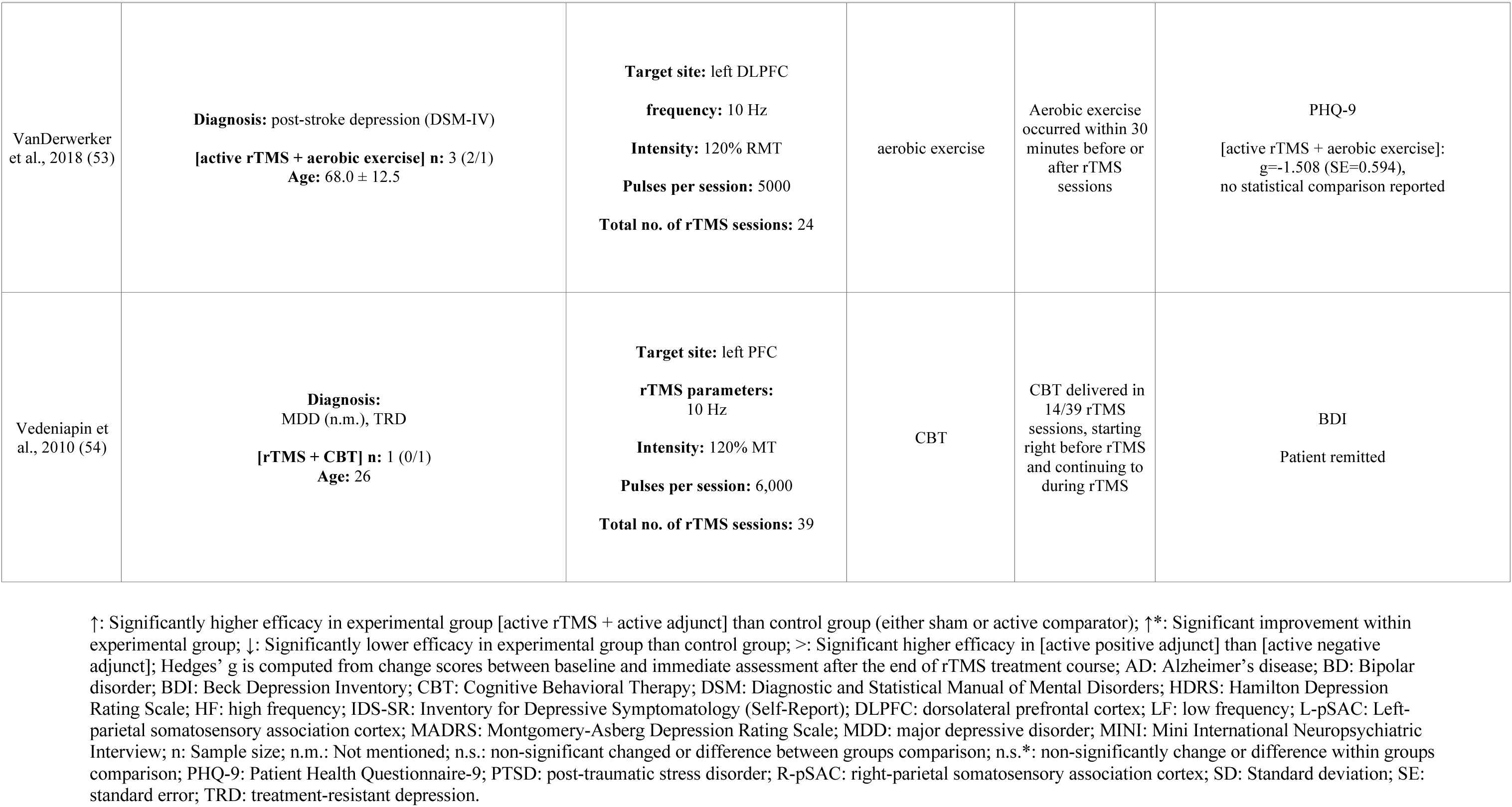

Thirteen of twenty studies recruited patients diagnosed with major depressive disorder (MDD), post-stroke depression, or persistent depressive disorder (dysthymia) (35, 38, 39, 41, 43–46, 48, 50, 51, 53, 54):. Six other studies recruited patients with other neuropsychiatric conditions with comorbid depressive symptoms, namely, Alzheimer’s disease (36), cocaine use disorder (47), and post-traumatic stress disorder (42, 49, 52). MDD patients in eight included studies were treatment resistant (35, 38, 39, 43, 46, 48, 51, 54), with one study also recruiting patients with treatment resistance in bipolar disorder (35). Schedules of psychological methods combined with rTMS is illustrated in **Figure 2**.

**Figure 2.**
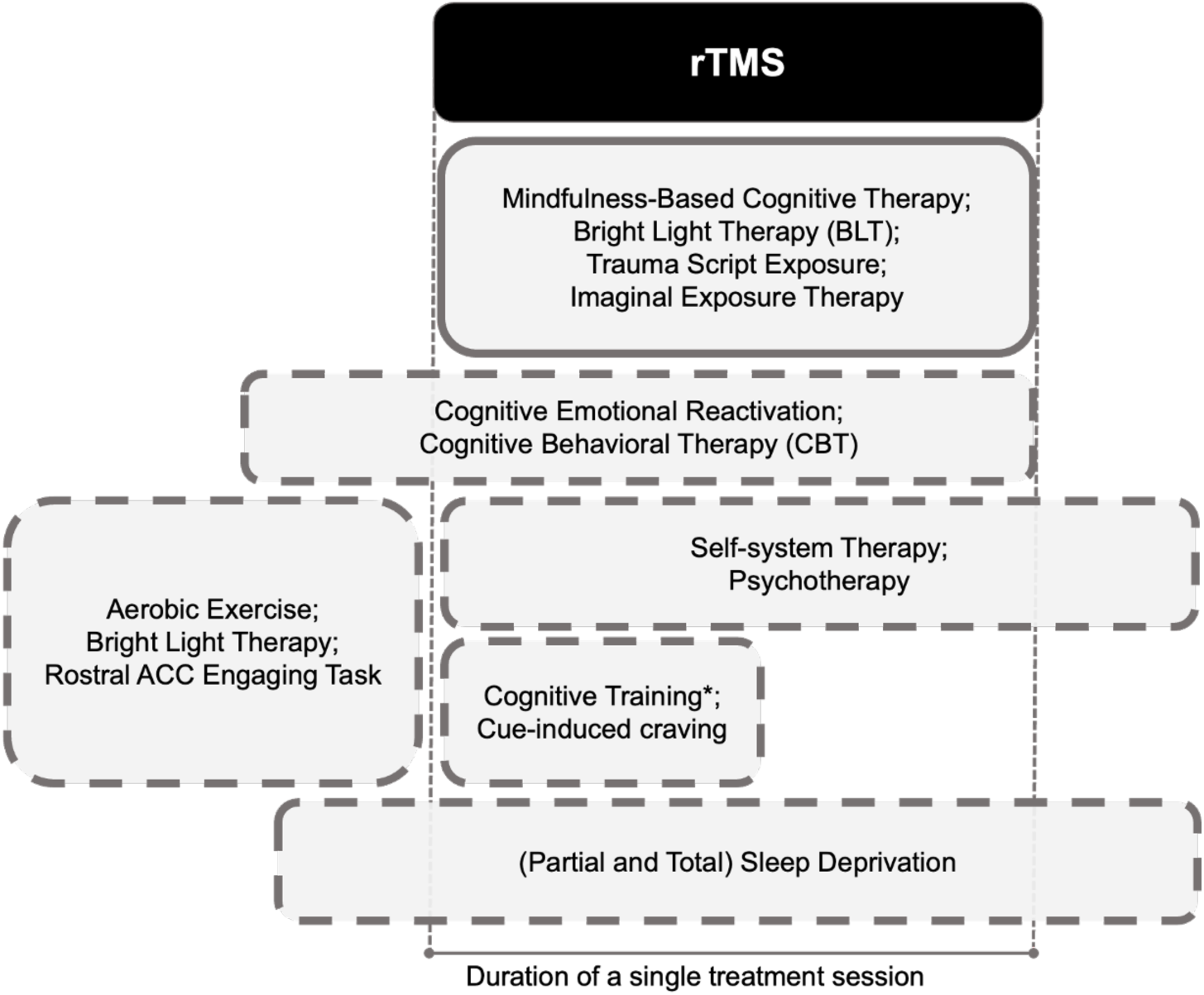
Boxes indicate schedules of psychological interventions that have been attempted before or during sessions of rTMS. For example, the first box below “rTMS” indicates complete concurrence; the next box indicates interventions that start before rTMS then continue through an rTMS session. Box dimensions are not scaled to durations or any property of the interventions (e.g., dose). *Cognitive training was interleaved with rTMS trains. Abbreviations: rTMS, repetitive transcranial magnetic stimulation; ACC, anterior cingulate cortex.

The DLPFC was a prime target, with seventeen of twenty studies targeting this region (35, 36, 38–52): sixteen targeted the left DLPFC (35, 36, 38–48, 50, 51, 53), two studies targeted the right DLPFC (49, 52), and four studies targeted bilateral DLPFC (36, 38, 39, 42).

### Meta-analysis results of depression change scores across disorders

The number of included studies allowed for three meta-analyses (of four discussed in *Groups and Comparisons* section above): within-group comparisons of [active rTMS + active PSYC], or endpoint minus baseline scores; between group comparisons of [active rTMS + active PSYC] versus [active rTMS + sham PSYC]; and between-group comparisons of [active rTMS + active PSYC] versus [sham rTMS + active PSYC].

Seventeen studies (20 datasets) reported endpoint versus baseline changes for an [active rTMS + active PSYC] treatment. This uncontrolled effect on depression severity across disorders was large and significantly therapeutic with a corrected standardized mean difference (g) of - 1.91, (standard error (SE) = 0.45, 95% confidence interval (CI) = −2.80 to −1.03), p < 0.01). However, these results were substantially heterogeneous (Q(df=19) = 126.59, p < 0.01, I² = 97.27%). Meta-regression results suggest age and sex ratio are non-significant moderators (p>0.05), but whether PSYC was an antidepressant intervention was significant (coefficient=-1.745, SE=0.819, 95%CI=-3.350 to −0.140, p=0.0331). This significant negative moderation indicates that depression severity decreases further when rTMS is combined with an intervention for depression. Leave-one-out sensitivity tests indicated a robust meta-analytic estimate, with g ranging between −2.01 to −1.41 (p < 0.01 for each iteration). Publication bias may possibly be an issue: despite Egger’s test being non-significant, the funnel plot appears asymmetric with bias towards lower efficacy (**Figure 3a**).

**Figure 3.**
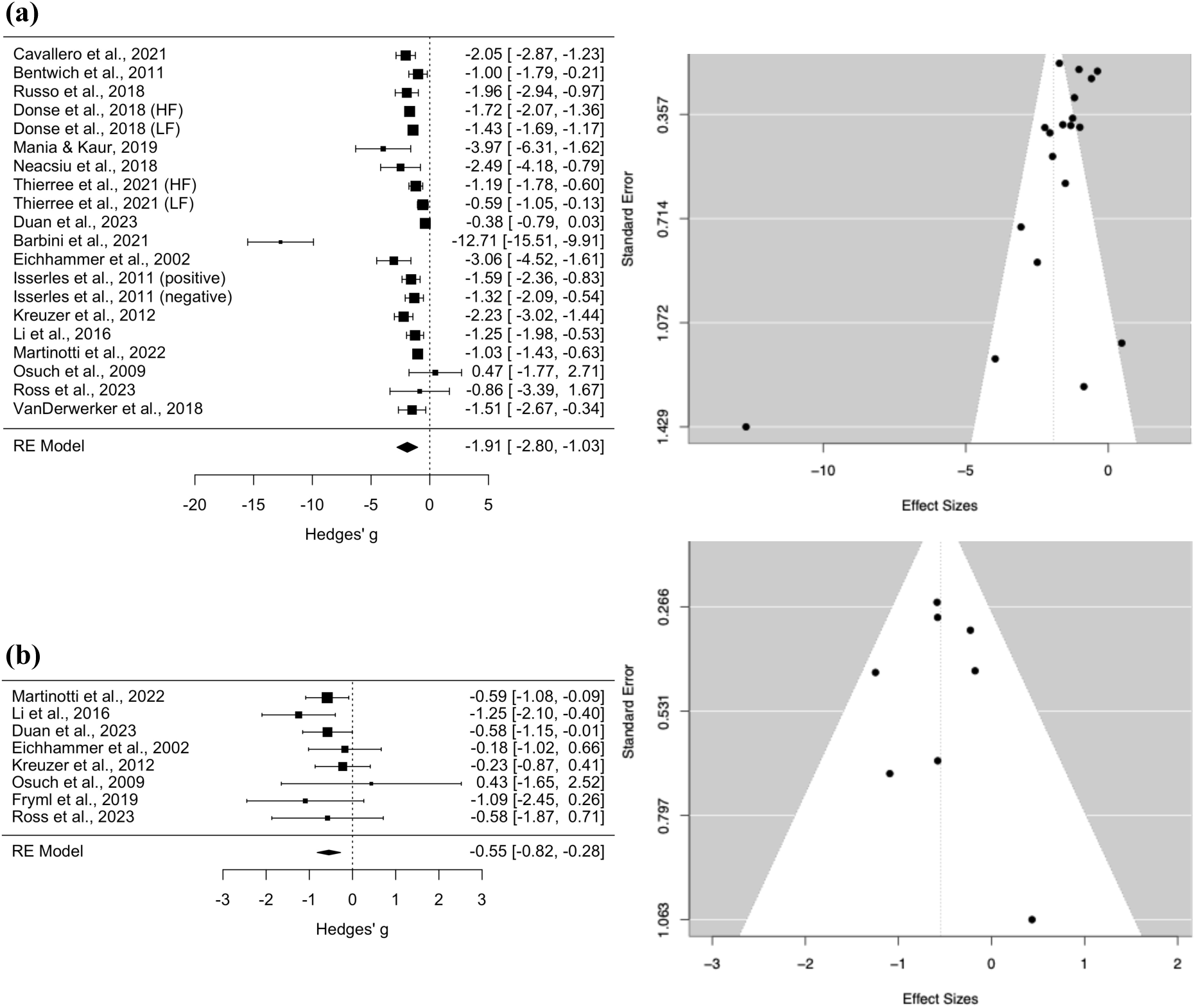
(a) Pooled within-group comparisons (endpoint - baseline) of [active rTMS + active PSYC] treatment. (b) Between-group comparisons of change scores between [active rTMS + active PSYC] and [sham rTMS + active PSYC] groups. Black box sizes indicate weight in the pooled estimate.

Eight studies compared [active rTMS + active PSYC] versus [sham rTMS + active PSYC]. This controlled effect on depression severity across disorders was medium and significantly therapeutic with g of −0.55 (SE = 0.14, 95% CI = −0.82 to −0.28, p < 0.01). These results were not heterogenous (Q(df=7)=5.84, p=0.56, I² = 0.00%). Leave-one-out sensitivity tests indicated a robust meta-analytic estimate, with g ranging between −0.62 to −0.47 (p < 0.01 for each iteration). Publication bias did not appear to be an issue as the funnel plot appears symmetric (**Figure 3b**).

Four studies compared groups receiving [active rTMS + active PSYC] versus [active rTMS + sham PSYC]. This controlled effect on depression severity was not significant. Pooled study effects were substantially heterogeneous (Q(df=3)=46.00, p<0.01, I² = 90.22%). Leave-one-out sensitivity tests indicated a poor reliability of the estimate, with g ranging between −0.32 to +0.44 (each iteration remained non-significant). The funnel plot appeared symmetric, indicating unlikely publication bias (**Supplementary Text 2**).

### Secondary meta-analysis: left DLPFC treatment for depressive disorders

The above meta-analyses were repeated but only including studies that targeted the left DLPFC to treat patients with depressive disorders (e.g., MDD or post-stroke depression). Forest and funnel plots are provided in **Supplementary Text 3**. Meta-analytic estimates are shown in **Figure 4**.

**Figure 4.**
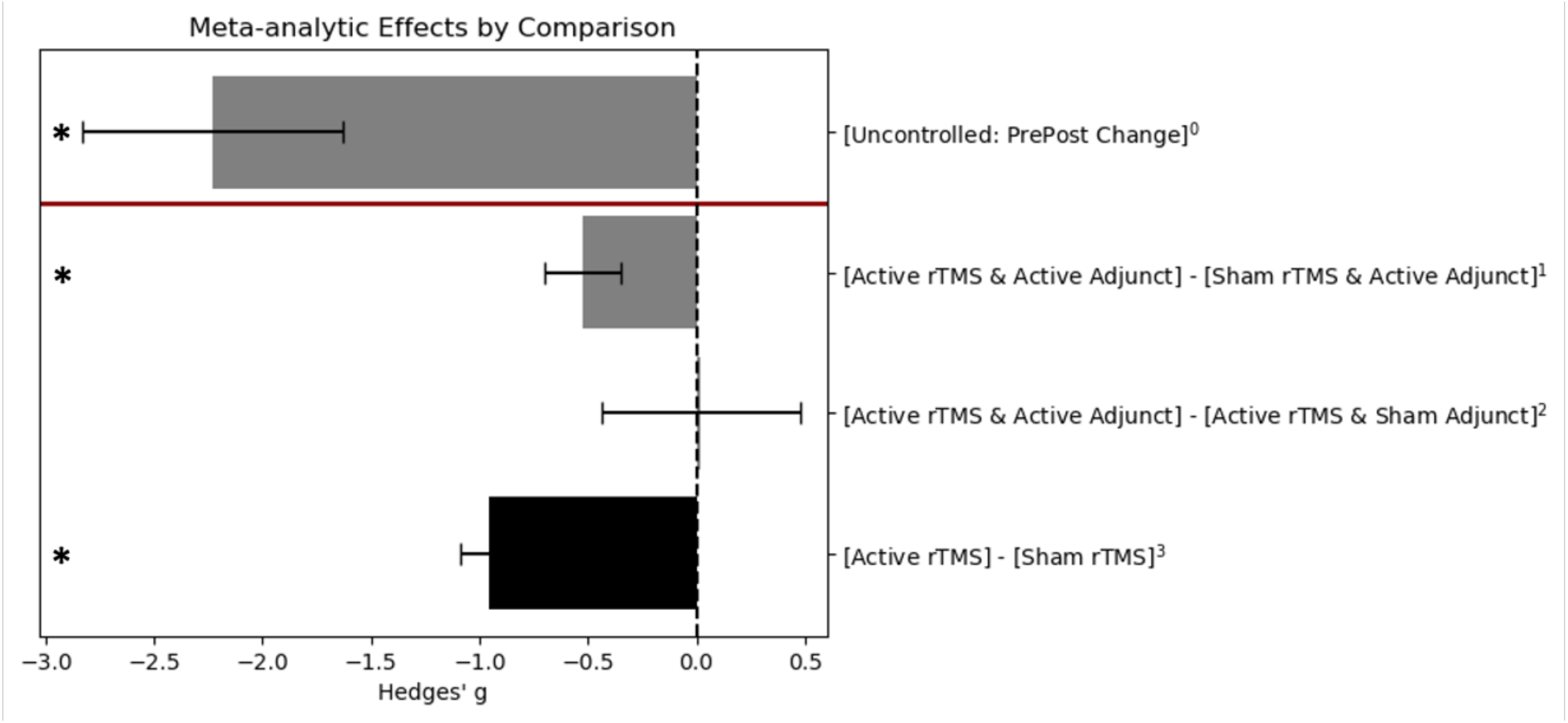
Meta-analytic estimates of studies treating patients with depressive disorders and targeting the left dorsolateral prefrontal cortex (DLPFC). Blue bars are based on findings in this review; the red bar is based on findings from Kan et al., Lancet Psychiatry (3). ^0^Estimate is based on 14 uncontrolled studies (16 datasets); ^1^estimate is based on five controlled trials; ^2^estimate is based on four controlled trials; ^3^estimate is based on 61 controlled trials. * Pooled estimate was statistically significant (p<0.01).

Within-group comparisons of [active rTMS + active PSYC] at endpoint minus baseline change scores included 14 studies (16 datasets), with a large and significantly therapeutic g of - 2.23, (SE = 0.60, 95% CI = −3.41 to −1.05), p < 0.01), albeit with substantial heterogeneity (Q(df=15) = 111.21, p < 0.01, I² = 97.74%). Mixed findings for publication bias were observed: Egger’s test was not significant, but the funnel plot was asymmetric. Leave-one-out sensitivity tests suggested robust findings (g-range = −2.39 to −1.59, p<0.01 for each iteration). Age, sex ratio, and whether PSYC was an antidepressant therapy were non-significant moderators.

Five studies compared [active rTMS + active PSYC] versus [sham rTMS + active PSYC] and targeted the left DLPFC of patients with depressive conditions. The effect was medium and significantly therapeutic with g of −0.52 (SE = 0.18, 95%CI = −0.87 to −0.18, p < 0.01). The results were not heterogenous (Q(df=4)=4.31, p=0.36, I² = 4.11%). However, leave-one-out sensitivity tests indicated an unstable meta-analytic estimate, with g ranging from −0.64 to −0.39, (only non-significant when Duan et al. (40) was excluded). Publication bias may not be an issue as the funnel plot appeared symmetric.

The fours studies that compared [active rTMS + active PSYC] versus [active rTMS + sham PSYC] groups all applied rTMS to the left DLPFC of patients with depressive disorders.

### Power analysis results

Power analysis curves are included in the **Supplementary Text 4**. Average change scores reported by Li et al. (45) were imputed for the following groups: [active rTMS + active PSYC], [active rTMS + sham PSYC], and [sham rTMS + active PSYC]. Li et al.’s study was selected as these authors included two active comparators and their findings supported the hypothesis that combining rTMS with PSYC improved antidepressant efficacy over either alone. However, Li et al. did not include a [sham rTMS + sham PSYC] group; we imputed average change scores for this group from Duan et al. (40), as these authors used the same standardized assessment (17-item Hamilton Depression Rating Scale), treated patients diagnosed with MDD, and combined sham rTMS with a control for active PSYC.

To test the hypothesis that [active rTMS + active PSYC] is significantly better than [sham rTMS + active PSYC] and [sham rTMS + sham PSYC], but not compared to [active rTMS + sham PSYC], a clinical trial with four independent groups would need to recruit 80 patients (20 patients per group). For the hypothesis that both active treatment is better than all active and sham comparators (including [active rTMS + sham PSYC]), the number of patients needed jumps to 240 (60 patients per group). This increased sample size requirement for sufficient power is due to the small difference in average change scores between the [active rTMS + active PSYC] and [active rTMS + sham PSYC] groups. Given these findings, only two included studies (35, 40) were sufficiently powered.

## Discussion

Following PRISMA-ScR guidelines (29), this scoping review and meta-analysis chartered the literature on the antidepressant efficacy of combining rTMS with psychological tasks or interventions. As of 10 July 2024, this literature comprised twenty studies, **Table 1**; (35–54), combining rTMS with psychotherapy, bright light therapy, aerobic exercise, partial and total sleep deprivation, cognitive training, cognitive emotional reactivation, and a psychophysical task.

Numerous systematic reviews and meta-analyses have investigated the antidepressant efficacy of pharmacological, psychological, and non-invasive brain stimulation therapies. For example, an umbrella review of meta-analyses found a small yet significant effect size when pharmacological or psychological treatments are used independently (55). The efficacy of rTMS alone also shows promise, with a meta-analysis of various rTMS protocols indicating small to medium effect sizes on depressive conditions (2). In contrast, a cross-diagnostic meta-analysis focusing on protocols targeting the left DLPFC found a medium to large effect size for depressive symptoms across neuropsychiatric conditions (3).

There is increasing interest in identifying synergies between treatments to advance these approaches. For instance, Rakesh et al. (56) recently conducted a meta-analysis on the synergistic effects of rTMS with pharmacological treatments, observing a large effect size of rTMS with antidepressants compared to sham rTMS with antidepressant. This scoping review contributes to the field by providing meta-analytic estimates of the efficacy of combining rTMS with psychological tasks and interventions. Results suggest that while the antidepressant efficacy of combining rTMS with psychological methods is promising, it does not surpass the efficacy of rTMS alone.

### Uncontrolled meta-analysis results

Uncontrolled clinical trials show a large, significant, and robust short-term reduction of depressive symptom severity, measured by comparing treatment endpoint to baseline scores. However, these results are substantially heterogenous, and publication bias is possible. The meta-analysis findings remain consistent even when limited to studies targeting the left DLPFC for treating depressive conditions, such as MDD, post-stroke depression, or comorbid MDD with PTSD or dysthymia. Meta-regressions showed that age and sex were not significant moderators for both pooled estimates. However, whether the concurrent psychological method treated depressive symptoms was a significant moderator for the pooled estimate across protocols and diagnoses (**Figure 3a**), but not significant when focusing on left DLPFC protocols for patients with depressive conditions (**Supplementary Text 3**). The difference may be due to highly mixed outcomes, as pooled estimates showed significant and substantial heterogeneity. Additionally, the notably high efficacy of left DLPFC protocols for depressive symptoms may also overshadow the antidepressant effects of psychological augmentations – a recent cross-diagnostic meta-analysis of 61 clinical trials observed a Hedge’s g of 0.959 (95%CI=-1.209 to - 0.708) with rTMS alone compared to sham rTMS alone (3). While the large effect sizes from the uncontrolled meta-analysis show promise and support further investigations, the estimates are uncontrolled and exhibit substantial heterogeneity.

### Controlled meta-analysis results

Control groups are essential baselines for distinguishing combination treatment effects from confounding variables like placebo effects and natural symptom fluctuations in episodic conditions such as depression. Additionally, to draw causal conclusions about synergistic, antagonistic, or null effects of psychological method combinations with rTMS, antidepressant outcomes must be compared with rTMS alone and psychological methods alone. For example, if combining rTMS with a psychological interventions result in a moderate effect size, but rTMS alone produces a large effect, the outcome might be misinterpreted as synergistic when it is actually antagonistic. Such antagonistic effects may explain findings by Isserles et al. (43). An additional concern for rTMS is that its antidepressant effects are substantially heterogeneous (3–5), making drawing conclusions from uncontrolled observations highly problematic. However, there are challenges with a control condition for psychological interventions, such that few studies could have included sham conditions.

Given these challenges, it was possible to pool study findings for two types of comparisons. In the first comparison, pooling results from seven studies, the active combination treatment [active rTMS + active PSYC] was significantly more effective than [sham rTMS + active PSYC], with a medium effect size. This robust meta-analytic estimate was consistent and supported by a funnel plot indicating no publication bias. In the second comparison, pooling results from three studies, the effect size for [active rTMS + active PSYC] versus [active rTMS + sham PSYC] was non-significant and highly heterogeneous. These results suggest that combining psychological methods with rTMS enhances efficacy, but the reverse effect is not supported. Although it must be emphasized that the number of pooled studies was small for both comparisons, e.g., multiple analyses requiring more than ten pooled studies not recommended.

The results of various meta-analytic estimates are summarized in **Figure 3a** and highlighted by a power analysis (**Supplementary Text 4**). The meta-analytic effect size of active rTMS compared to sham rTMS in patients with depressive conditions is significantly large, though notably heterogeneous (3). Relative to this effect size, the benefits of including a psychological task or intervention to enhance the efficacy of rTMS is not supported. The simulations-based power analysis highlights this difference in effect size. Using the observed effect sizes of [active rTMS + active PSYC] versus rTMS alone, and assuming significant superiority of combination over rTMS alone, the number of patients needed to obtain sufficient power is 80 patients per group (240 total). Compared with 20 patients per group (80 total) if simply testing the hypothesis that the combination treatment is better that sham or the psychological method alone.

### Notable observations from included studies

Bright light therapy with rTMS is the most promising combination therapy for depression. Two controlled studies reported encouraging outcomes: a large effect size indicated superior efficacy when bright light therapy was administered on mornings of rTMS treatment days and compared to rTMS alone (35); and a separate research team observed large reductions of HDRS scores in six patients with treatment resistant depression when bright light therapy was administered during rTMS sessions (46).

This scoping review was motivated by concerns that brain states may influence the antidepressant effects of rTMS. Two identified studies illustrate these concerns through controlled designs. During rTMS, Isserless et al. (43) guided patients towards negative or positive thinking during rTMS, operationalized as thoughts that mitigate or exacerbate depressive symptoms. Treatment outcomes for these groups were compared to patients receiving rTMS alone. Assessed using Beck’s Depression Index, the authors identified a significant antagonistic effect by the negative thinking condition. These findings need to be replicated as the sample size was small, the finding was not observed when assessing symptom severity with the HDRS, and the design of Isserles et al. (43) may not dissociate whether negative thinking blocked the effects of rTMS (a ‘brain state’ effect) or reinforced depressive symptoms. Nevertheless, the authors cautioned the need to control for brain state during antidepressant protocols of rTMS (43). In contrast, Li et al. (45) had patients complete a shape-discrimination task just before high frequency rTMS, as this task was observed to induce frontal electroencephalogram theta activity (57), which persisted after the completion of the psychological task (45). This combination significantly boosted antidepressant effects compared to [active rTMS + sham PSYC] (with a small effect size;**Table 2**).

### Limitations

Our screening protocol by title and abstract may have missed studies if these details were only provided in the main text and not the title or abstract; or if they were not discussed in relevant reviews (24–27) or not in the reference list of fully screened studies. Meta-analyses for controlled studies were also based on a small number of studies, restricting our analysis to pooled statistics and funnel plots only; Egger’s tests or meta-regressions when study count was below ten were not conducted due to limited power of such analyses.

## Conclusion

Notwithstanding the low number of studies, when comparing the meta-analytic findings of combined treatments (**Figure 4**) with literature on rTMS alone (2, 3), where patients are typically instructed to relax during sessions, rTMS shows promise as an augmentation for psychological interventions. However, for rTMS alone, simply instructing patients to relax seems sufficient for effective antidepressant protocols.

## Data Availability

All data produced in the present study are available upon reasonable request to the authors.

## Author contributions

Conceptualization and funding, CGG, GSK; literature screening, CGG, YTC, HYCC, SWC, YYC, AHPT, MJ; chartering table, CGG; data extraction, CGG; extracted data verification, MX and AHPT; data analysis, CGG; data interpretation, CGG and GSK; writing first draft, CGG; revisions to draft, AHPT, MJ, GSK.

## Acknowledgements

We would like to thank our former students for assisting in the initial literature screening: Yuet Tung Chan (YTC), Chui Yee Cheung (CYC), Hoi Yan Clarice Cheuk (HYCC), Sze Wing Cheung, (SWC) and Ying Yau Choi (YYC).

## Financial support

This work was supported by the General Research Fund under the University Grants Committee of the Hong Kong Special Administrative Region (numbers 15100120, 25100219 and 15106222) and the Mental Health Research Center, The Hong Kong Polytechnic University.

## Conflict of interest

None.

## Notes

### Competing Interest Statement

The authors have declared no competing interest.

### Clinical Protocols

https://osf.io/n8hw3

### Author Declarations

The scoping review and meta-analysis used ONLY openly available human data that were originally reported by published journal articles of included clinical trials that were approved by their respective Ethics Committee or Institutional Review Board (IRB).

