## Supplementary Materials for "Antidepressant efficacy of administering repetitive transcranial magnetic stimulation (rTMS) concurrently with psychological tasks or interventions: a scoping review and meta-analysis"

### CONTENTS


#### SUPPLEMENTARY TABLES 3

**Supplementary Table S1. Search queries (up-to 10 July 2024).** 3

**Supplementary Table S2. Checklist of the Reporting Items for Systematic reviews and Meta-Analyses extension for Scoping Reviews (PRISMA-ScR)** 4

**Supplementary Table S3. Studies excluded during full-text screening.** 6
*References by Motivation for Exclusion* 8

#### SUPPLEMENTARY TEXT 20

**Supplementary Text 1. *Effect sizes based on change scores of within-group and between-group comparisons*** 20
*Controlled Effect Sizes* 21
*Uncontrolled Effect Sizes* 22
*Special Cases* 23

**Supplementary Text 2. *Meta-analysis results of depression change scores across disorders*** 24

**Supplementary Text 3: *Secondary meta-analysis: left DLPFC treatment for depressive disorders*** 25

**Supplementary Text 4: *Power analysis methods and results*** 26

#### Supplementary Tables

**Supplementary Table S1.** Search queries (up-to 10 July 2024).

| |
|---|
| PubMed:<br><br>(“transcranial magnetic stimulation"[Title/Abstract] OR TMS[Title/Abstract] OR rTMS[Title/Abstract] OR "repetitive transcranial magnetic stimulation"[Title/Abstract] OR "theta burst stimulation"[Title/Abstract] OR TBS[Title/Abstract] OR "intermittent theta burst stimulation"[Title/Abstract] OR iTBS[Title/Abstract] OR "continuous theta burst stimulation"[Title/Abstract] OR cTBS[Title/Abstract] OR "deep transcranial magnetic stimulation"[Title/Abstract] OR dTBS[Title/Abstract]) AND (depression[Title/Abstract] OR depressive[Title/Abstract] OR MDD[Title/Abstract] OR Treatment resistant depression[Title/Abstract] OR TRD[Title/Abstract]) |
| Web of Science; filters set to search titles and abstracts:<br><br>(“transcranial magnetic stimulation" OR TMS OR rTMS OR "repetitive transcranial magnetic stimulation" OR "theta burst stimulation" OR TBS OR "intermittent theta burst stimulation" OR iTBS OR "continuous theta burst stimulation" OR cTBS OR "deep transcranial magnetic stimulation" OR dTBS) AND (depression OR depressive OR MDD OR Treatment resistant depression OR TRD) |

**Supplementary Table S2.** Checklist of the Reporting Items for Systematic reviews and Meta-Analyses extension for Scoping Reviews (PRISMA-ScR)

| SECTION | ITEM | PRISMA-ScR CHECKLIST ITEM | REPORTED ON PAGE # |
|---|---|---|---|
| **TITLE** | | | |
| Title | 1 | Identify the report as a scoping review. | 1 |
| **ABSTRACT** | | | |
| Structured summary | 2 | Provide a structured summary that includes (as applicable): background, objectives, eligibility criteria, sources of evidence, charting methods, results, and conclusions that relate to the review questions and objectives. | 1 |
| **INTRODUCTION** | | | |
| Rationale | 3 | Describe the rationale for the review in the context of what is already known. Explain why the review questions/objectives lend themselves to a scoping review approach. | 4-5 |
| Objectives | 4 | Provide an explicit statement of the questions and objectives being addressed with reference to their key elements (e.g., population or participants, concepts, and context) or other relevant key elements used to conceptualize the review questions and/or objectives. | 5 |
| **METHODS** | | | |
| Protocol and registration | 5 | Indicate whether a review protocol exists; state if and where it can be accessed (e.g., a Web address); and if available, provide registration information, including the registration number. | 5 |
| Eligibility criteria | 6 | Specify characteristics of the sources of evidence used as eligibility criteria (e.g., years considered, language, and publication status), and provide a rationale. | 6 |
| Information sources* | 7 | Describe all information sources in the search (e.g., databases with dates of coverage and contact with authors to identify additional sources), as well as the date the most recent search was executed. | 6-7 |
| Search | 8 | Present the full electronic search strategy for at least 1 database, including any limits used, such that it could be repeated. | Supp. Table 1 |
| Selection of sources of evidence† | 9 | State the process for selecting sources of evidence (i.e., screening and eligibility) included in the scoping review. | 7 |
| Data charting process‡ | 10 | Describe the methods of charting data from the included sources of evidence (e.g., calibrated forms or forms that have been tested by the team before their use, and whether data charting was done independently or in duplicate) and any processes for obtaining and confirming data from investigators. | 7 |
| Data items | 11 | List and define all variables for which data were sought and any assumptions and simplifications made. | 7 |
| Critical appraisal of individual sources of evidence§ | 12 | If done, provide a rationale for conducting a critical appraisal of included sources of evidence; describe the methods used and how this information was used in any data synthesis (if appropriate). | n/a |

| SECTION | ITEM | PRISMA-ScR CHECKLIST ITEM | REPORTED ON PAGE # |
|---|---|---|---|
| Synthesis of results | 13 | Describe the methods of handling and summarizing the data that were charted. | 7-9 |
| **RESULTS** | | | |
| Selection of sources of evidence | 14 | Give numbers of sources of evidence screened, assessed for eligibility, and included in the review, with reasons for exclusions at each stage, ideally using a flow diagram. | 10 |
| Characteristics of sources of evidence | 15 | For each source of evidence, present characteristics for which data were charted and provide the citations. | 11 |
| Critical appraisal within sources of evidence | 16 | If done, present data on critical appraisal of included sources of evidence (see item 12). | n/a |
| Results of individual sources of evidence | 17 | For each included source of evidence, present the relevant data that were charted that relate to the review questions and objectives. | 12-21 |
| Synthesis of results | 18 | Summarize and/or present the charting results as they relate to the review questions and objectives. | 22-24 |
| **DISCUSSION** | | | |
| Summary of evidence | 19 | Summarize the main results (including an overview of concepts, themes, and types of evidence available), link to the review questions and objectives, and consider the relevance to key groups. | 25-27 |
| Limitations | 20 | Discuss the limitations of the scoping review process. | 27 |
| Conclusions | 21 | Provide a general interpretation of the results with respect to the review questions and objectives, as well as potential implications and/or next steps. | 27 |
| **FUNDING** | | | |
| Funding | 22 | Describe sources of funding for the included sources of evidence, as well as sources of funding for the scoping review. Describe the role of the funders of the scoping review. | 27 |

JBI = Joanna Briggs Institute; PRISMA-ScR = Preferred Reporting Items for Systematic reviews and Meta-Analyses extension for Scoping Reviews.
* Where *sources of evidence* (see second footnote) are compiled from, such as bibliographic databases, social media platforms, and Web sites.
† A more inclusive/heterogeneous term used to account for the different types of evidence or data sources (e.g., quantitative and/or qualitative research, expert opinion, and policy documents) that may be eligible in a scoping review as opposed to only studies. This is not to be confused with *information sources* (see first footnote).
‡ The frameworks by Arksey and O'Malley (6) and Levac and colleagues (7) and the JBI guidance (4, 5) refer to the process of data extraction in a scoping review as data charting.
§ The process of systematically examining research evidence to assess its validity, results, and relevance before using it to inform a decision. This term is used for items 12 and 19 instead of "risk of bias" (which is more applicable to systematic reviews of interventions) to include and acknowledge the various sources of evidence that may be used in a scoping review (e.g., quantitative and/or qualitative research, expert opinion, and policy document).

*From:* Tricco AC, Lillie E, Zarin W, O'Brien KK, Colquhoun H, Levac D, et al. PRISMA Extension for Scoping Reviews (PRISMAScR): Checklist and Explanation. Ann Intern Med. 2018;169:467–473. doi: 10.7326/M18-0850.

**Supplementary Table S3.** Studies excluded during full-text screening.

| Motivation for Exclusion | Study | Number of studies |
|---|---|---|
| rTMS did not include or was not concurrent with a psychological task or intervention | Adams et al., 2014; Adu et al., 2023; Albuquerque et al., 2018; Arns et al., 2012; Bech et al., 2014; Berlim et al., 2011; Bi et al., 2024; Borckardt et al., 2008; Bovy et al., 2019; Bröcker et al., 2019; Bruno et al., 2021; Charnsil et al., 2012; Chen et al., 2013; Chen et al., 2022a; Chen et al., 2022b; Concerto et al., 2015; Dai et al., 2023; Dan et al., 2023; Del Felice et al., 2016; Deschamps et al., 2016; Deschamps et al., 2018; Di Ponzio et al., 2023; Formánek et al., 2018; Frank et al., 2011; Frick et al., 2021; Fujita & Koga, 2005; Garg et al., 2016; Griffiths et al., 2022; Guinot et al., 2021; Hansen et al., 2004; Hausmann et al., 2004[a]; Hausmann et al., 2004[b]; Hermann et al., 2017; Hoflich et al., 1993; Hu et al., 2022; Huang et al., 2005; Huang et al., 2022a; Huang et al., 2022b; Iliceto et al., 2018; Iznak et al., 2015; Khedr et al., 2014; Kozel et al., 2018; Krstić et al., 2014; Krstić & Ilić, 2014; Kullakçi & Sonkaya, 2021; Langguth et al., 2007; Leblhuber et al., 2019; Leblhuber et al., 2021; Lee et al., 2019; Lee et al., 2016; Leong et al., 2020; Li et al., 2004; Li et al., 2010; Li et al., 2022a; Li et al., 2022b; Li et al., 2022c; Li et al., 2021a; Li et al., 2021b; Li et al., 2023; Lin et al., 2023; Liu et al., 2022; Liu et al., 2024; Majdi et al., 2021; Mansur et al., 2011; Mayer et al., 2021; Mogg et al., 2008; Mollica et al., 2024; Niimi et al., 2020a; Niimi et al., 2020b; Ning et al., 2022; Nongpiur et al., 2011; Norred et al., 2021; Novák et al., 2024;Oh & Kim, 2011; Padberg et al., 2002; Pantazatos et al., 2023; Picarelli et al., 2010; Plewnia et al., 2014; Prasser et al., 2015; Rapinesi et al., 2013; Rapinesi et al., 2016; Reddy et al., 2022; Rich et al., 2016; Richter et al., 2017; Rodriguez et al., 2020; Rossini et al., 2005; Rotharmel et al. 2021; Ryan et al., 2022; Sampson et al., 2011; Sarkhel et al., 2010; Senda et al., 2023; Seo et al., 2016; Schulze et al., 2016; Schutter et al., 2010; Sharma et al., 2020; Shere et al., 2021; Singh et al., 2020; Song et al., 2024; Spronk et al., 2008; Su et al., 2005; Sureshkumar et al., 2014; Tadayonnejad et al., 2022; Taib et al., 2019; Tan et al., 2015; Tang et al., 2022; Tavares et al., 2017; Tavares et al., 2020; Tavarez et al., 2021; Teti Mayer et al., 2021; Tilbor et al., 2024; Tong et al., 2021; Torres et al., 2015; Uygur et al., 2024; Vaithianathan et al., 2022; Verma et al., 2018; Vidya et al., 2022; Wang et al., 2022; Wobrock et al., 2015; Yagci et al., 2014; Yamazako et al., 2022; Yan et al., 2023; Yu et al., 2022; Yuan et al., 2020; Zangen et al., 2021; Zavorotnyy et al., 2020; | 131 |

| | | |
|---|---|---|
| | Zendjidjian et al., 2014; Zhang et al., 2019; Zhang et al., 2020; Zhang et al., 2020; Zhang et al., 2021a; Zhang et al., 2021b | |
| Is protocol, conference paper, book chapter without new data, secondary analysis, or abstract | Abou El-Magd et al., 2020; Adu et al., 2023; Bulteau et al., 2020; Canali et al., 2012; Dalhuisen et al., 2022; Davis et al., 2023; Demitrack et al., 2009; Ekpo et al., 2023; Haque & Malik, 2017; He et al., 2011[b]; Hill et al., 2021; Hu & Lisanby, 2015; Iseger et al., 2018; Jha et al., 2017; Jin et al., 2011; Kujovik et al., 2024; Lee et al., 2014; Lee et al., 2019; Lee et al., 2006; Lee et al., 2009; Leuchter et al., 2014; Leuchter & Hunger, 2016; Maslenikov et al., 2019; Oathes et al., 2022; Stikhina et al., 1999; Tang et al., 2018; Tynan et al., 2023; Uribe et al., 2020; Vaishnavi & Brammer, 2022; Walpoth et al., 2003; Wen et al., 2022 | 31 |
| Depressive symptom severity not assessed | Carmi et al., 2018; Chen et al., 2021; Rabey et al., 2013; Ross et al., 2018; Sokhadze et al., 2014 | 5 |
| rTMS was not applied | Canali et al., 2014; Kaneko et al., 2024; Lissemore et al., 2019; Ross et al., 2024; Salehinejad et al., 2022 | 5 |
| Specialized NIBS protocol (e.g., bilateral protocols or EEG-guided), but not combined with psychological task or intervention | Kavakbasi et al., 2024; Leuchter et al., 2014; Li et al., 2020; McDonald et al., 2006; Price et al., 2010; Robertson & Mortimer, 2022; Sreepada et al., 2020; Zrenner et al., 2020 | 8 |
| Healthy participants recruited only | Dalhuisen et al., 2023; Grosshageur et al., 2024 | 2 |
| Could not retrieve full text | Yin et al., 2022 | 1 |
| | **Total:** | **183** |

*References by Motivation for Exclusion*

***rTMS did not include or was not concurrent with a psychological task or intervention***

Adams et al. Integration of cortical brain stimulation and exposure and response prevention for obsessive-compulsive disorder (OCD). Brain Stimul. 2014; 7(5):764-5.

Adu et al. Apparent Lack of Benefit of Combining Repetitive Transcranial Magnetic Stimulation with Internet-Delivered Cognitive Behavior Therapy for the Treatment of Resistant Depression: Patient-Centered Randomized Controlled Pilot Trial. Brain Sci. 2023; Feb 9;13(2):293.

Albuquerque et al. Effects of repetitive transcranial magnetic stimulation and trans-spinal direct current stimulation associated with treadmill exercise in spinal cord and cortical excitability of healthy subjects: A triple-blind, randomized and sham-controlled study. PLoS One. 2018;13(3):e0195276.

Arns et al. Neurophysiological predictors of non-response to rTMS in depression. Brain Stimul. 2012;5(4):569-76.

Bech et al. Psychometric analysis of the Melancholia Scale in trials with non-pharmacological augmentation of patients with therapy-resistant depression. Acta Neuropsychiatr. 2014;26(3):155-60.

Berlim et al. High frequency repetitive transcranial magnetic stimulation as an augmenting strategy in severe treatment-resistant major depression: a prospective 4-week naturalistic trial. J Affect Disord. 2011;130(1-2):312-7.

Bi et al. Brain stimulation over the left DLPFC enhances motivation for effortful rewards in patients with major depressive disorder. Journal of Affective Disorders. 2024; 356: p. 414-423.

Borckardt et al. Significant analgesic effects of one session of postoperative left prefrontal cortex repetitive transcranial magnetic stimulation: a replication study. Brain Stimul. 2008;1(2):122-7.

Bovy et al. Combining attentional bias modification with dorsolateral prefrontal rTMS does not attenuate maladaptive attentional processing. Sci Rep. 2019;9(1):1168.

Bröcker et al. Accelerated theta-burst repetitive transcranial magnetic stimulation for depression in South Africa. S Afr J Psychiatr. 2019;25(0):1346.

Bruno et al. Combining intensive repetitive transcranial magnetic stimulation with neurofeedback in a case of treatment-resistant depression. Indian J Psychiatry. 2021;63(2):199-200.

Charnsil et al. An open-label study of adjunctive repetitive transcranial magnetic stimulation (rTMS) for partial remission in major depressive disorder. Int J Psychiatry Clin Pract. 2012;16(2):98-102.

Chen et al. Superior antidepressant effect occurring 1 month after rTMS: add-on rTMS for subjects with medication-resistant depression. Neuropsychiatr Dis Treat. 2013;9:397-401.

Chen et al. Comparative Analysis of the Effects of Escitalopram, Pramipexole, and Transcranial Magnetic Stimulation on Depression in Patients With Parkinson Disease: An Open-Label Randomized Controlled Trial. Clinical Neuropharmacology 2022a; 45(4):84-88.

Chen et al. High-frequency repetitive transcranial magnetic stimulation alleviates the cognitive side effects of electroconvulsive therapy in major depression. Front Psychiatry. 2022b; 13:1002809.

Concerto et al. Repetitive transcranial magnetic stimulation in patients with drug-resistant major depression: A six-month clinical follow-up study. Int J Psychiatry Clin Pract. 2015;19(4):252-8.

Dai et al. The Effect of Continuous Theta Burst Stimulation over the Right Dorsolateral Prefrontal Cortex on Cognitive Function and Emotional Regulation in Patients with Cerebral Small Vessel Disease. Brain Sci. 2023; 13(9).

Dan et al. Effect of Recurrent TMS and psychological therapy on Parkinson's Patient's cognitive performance, anxiety, and depressed mood. Archives of Clinical Psychiatry. 2023; 50(1), 57-62.

Del Felice et al. Neurophysiological, psychological and behavioural correlates of rTMS treatment in alcohol dependence. Drug Alcohol Depend. 2016;158:147-53.

Deschamps et al., Dynamics of postural control during repetitive transcranial magnetic stimulation in an adult with major depressive disorder. Aust N Z J Psychiatry, 2018: p. 291-293.

Deschamps et al. Posture cognitive dual-tasking: A relevant marker of depression-related psychomotor retardation. An illustration of the positive impact of repetitive transcranial magnetic stimulation in patients with major depressive disorder. J Psychiatr Res. 2016;83:86-93.

Di Ponzio et al. rTMS investigation of resistant Obsessive-Compulsive Related Disorders: Efficacy of targeting the reward system. Front Psychiatry. 2023 Feb 3;13:1035469.

Formánek et al. Combined transcranial magnetic stimulation in the treatment of chronic tinnitus. Ann Clin Transl Neurol. 2018;8;5(7):857-864.

Frank et al. Transcranial magnetic stimulation for the treatment of depression: feasibility and results under naturalistic conditions: a retrospective analysis. Eur Arch Psychiatry Clin Neurosci. 2011;261(4):261-6.

Frick et al. Habitual caffeine consumption moderates the antidepressant effect of dorsomedial intermittent theta-burst transcranial magnetic stimulation. J Psychopharmacol. 2021;35(12):1536-41.

Fujita & Koga. Clinical application of single-pulse transcranial magnetic stimulation for the treatment of depression. Psychiatry Clin Neurosci. 2005;59(4):425-32.

Garg et al. The efficacy of cerebellar vermal deep high frequency (theta range) repetitive transcranial magnetic stimulation (rTMS) in schizophrenia: A randomized rater blind-sham controlled study. Psychiatry Res. 2016;243:413-20.

Griffiths et al. Treatment resistant depression (TRD) service outpatient's experience of sleep, activity, and using a Fitbit wearable activity and sleep tracker. Mental Health Review Journal 2022; 27(2):158-174.

Guinot et al. Effects of Repetitive Transcranial Magnetic Stimulation and Multicomponent Therapy in Patients With Fibromyalgia: A Randomized Controlled Trial. Arthritis Care Res (Hoboken). 2021;73(3):449-58.

Hansen et al. Repetitive transcranial magnetic stimulation as add-on antidepressant treatment. The applicability of the method in a clinical setting. Nord J Psychiatry. 2004;58(6):455-7.

Hausmann et al. No benefit derived from repetitive transcranial magnetic stimulation in depression: a prospective, single centre, randomised, double blind, sham controlled "add on" trial. J Neurol Neurosurg Psychiatry. 2004[a];75(2):320-2.

Hausmann et al. No deterioration of cognitive performance in an aggressive unilateral and bilateral antidepressant rTMS add-on trial. J Clin Psychiatry. 2004[b];65(6):772-82.

Herrmann et al. Medial prefrontal cortex stimulation accelerates therapy response of exposure therapy in acrophobia. Brain Stimul. 2017;10(2):291-7.

Hoflich et al. APPLICATION OF TRANSCRANIAL MAGNETIC STIMULATION IN TREATMENT OF DRUG-RESISTANT MAJOR DEPRESSION - A REPORT OF 2 CASES. Human Psychopharmacology-Clinical and Experimental 1993; 8(5): 361-365.

Hu et al. Repetitive transcranial magnetic stimulation combined with cognitive behavioral therapy treatment in alcohol-dependent patients: A randomized, double-blind sham-controlled multicenter clinical trial. Front Psychiatry. 2022;13:935491.

Huang et al. Repetitive transcranial magnetic stimulation for treating medication-resistant depression in Taiwan: a preliminary study. J Chin Med Assoc. 2005;68(5):210-5.

Huang et al. An Exploratory Study of a Novel Combined Therapeutic Modality for Obsessive-Compulsive Disorder. Brain Sci. 2022;12(10):1309. (a)

Huang et al. Increased Prefrontal Activation During Verbal Fluency Task After Repetitive Transcranial Magnetic Stimulation Treatment in Depression: A Functional Near-Infrared Spectroscopy Study. Front Psychiatry. 2022; 13: 876136. (b)

Iliceto et al. Repetitive Transcranial Magnetic Stimulation for Treatment of Depression in a Patient With Severe Traumatic Brain Injury. Ochsner J. 2018;18(3):264-7.

Iznak et al. [Transcranial Magnetic Stimulation in the Combined Treatment of Pharmacoresistant Depression: Dynamics of Clinical, Psychological and EEG Parameters]. Fiziol Cheloveka. 2015;41(5):57-65.

Khedr et al. Dual-hemisphere repetitive transcranial magnetic stimulation for rehabilitation of poststroke aphasia: a randomized, double-blind clinical trial. Neurorehabil Neural Repair. 2014;28(8):740-50.

Kozel et al. Repetitive TMS to augment cognitive processing therapy in combat veterans of recent conflicts with PTSD: A randomized clinical trial. J Affect Disord. 2018;229:506-14.

Krstić et al. Low-frequency repetitive transcranial magnetic stimulation in the right prefrontal cortex combined with partial sleep deprivation in treatment-resistant depression: a randomized sham-controlled trial. J ECT. 2014;30(4):325-31.

Krstić & Ilić. Switch to hypomania induced by repetitive transcranial magnetic stimulation and partial sleep deprivation added to antidepressant: a case report. Vojnosanit Pregl. 2014;71(2):207-10.

Kullakçi & Sonkaya. The Investigation of the Effects of Repetitive Transcranialmagnetic Stimulation Treatment on Taste and Smell Sensations in Depressed Patients. Noro Psikiyatr Ars. 2021;58(1):26-33.

Langguth et al. Pre-treatment anterior cingulate activity as a predictor of antidepressant response to repetitive transcranial magnetic stimulation (rTMS). Neuro Endocrinol Lett. 2007;28(5):633-8.

Leblhuber et al. Treatment of patients with geriatric depression with repetitive transcranial magnetic stimulation. J Neural Transm (Vienna). 2019;126(8):1105-10.

Leblhuber et al. Repetitive transcranial magnetic stimulation in the treatment of resistant depression: changes of specific neurotransmitter precursor amino acids. J Neural Transm (Vienna). 2021;128(8):1225-31.

Lee et al. Effects of Repetitive Transcranial Magnetic Stimulation on Psychological Aspects of Patients with Chronic Lower Back Pain. J Magn. 2019;24(3): 543-548.

Lee et al. Treatment of Alzheimer's Disease with Repetitive Transcranial Magnetic Stimulation Combined with Cognitive Training: A Prospective, Randomized, Double-Blind, Placebo-Controlled Study. J Clin Neurol. 2016;12(1):57-64.

Leong et al. A Randomized Sham-controlled Trial of 1-Hz and 10-Hz Repetitive Transcranial Magnetic Stimulation (rTMS) of the Right Dorsolateral Prefrontal Cortex in Civilian Post-traumatic Stress Disorder: Un essai randomisé contrôlé simulé de stimulation magnétique transcrânienne repetitive (SMTr) de 1 Hz et 10 Hz du cortex préfrontal dorsolatéral droit dans le trouble de stress post-traumatique chez des civils. Can J Psychiatry. 2020 Nov;65(11):770-778.

Li et al. Can left prefrontal rTMS be used as a maintenance treatment for bipolar depression? Depress Anxiety. 2004;20(2):98-100.

Li et al. Antidepressant mechanism of add-on repetitive transcranial magnetic stimulation in medication-resistant depression using cerebral glucose metabolism. J Affect Disord. 2010;127(1-3):219-29.

Li et al. Factors associated with antidepressant responses to repetitive transcranial magnetic stimulation in antidepressant-resistant depression. Front Neurosci. 2022; Dec 2;16:1046920. (a)

Li et al. Mechanisms of Repetitive Transcranial Magnetic Stimulation on Post-stroke Depression: A Resting-State Functional Magnetic Resonance Imaging Study. Brain Topogr 2022; 35(3): 363-374. (b)

Li et al. Effect of Ipsilateral, Contralateral or Bilateral Repetitive Transcranial Magnetic Stimulation in Patients with Lateralized Tinnitus: A Placebo-Controlled Randomized Study. Brain Sci 2022; 12(6). (c)

Li et al. Co-activation patterns across multiple tasks reveal robust anti-correlated functional networks. Neuroimage 2021; 227. (a)

Li et al. Effectiveness and safety of repetitive transcranial magnetic stimulation for the treatment of morphine dependence: A retrospective study. Medicine (Baltimore). 2021;100(14):e25208. (b)

Li et al. Add-On Intermittent Theta Burst Stimulation Improves the Efficacy of First-Episode and Recurrent Major Depressive Disorder: Real-World Clinical Practice. Neuropsychiatric Disease and Treatment. 2023; 19: p. 109-116.

Lin et al. Effect of low-frequency repetitive transcranial magnetic stimulation as adjunctive treatment for insomnia patients under hypnotics: A randomized, double-blind, sham-controlled study. J Chin Med Assoc. 2023 Jun 1;86(6):606-613.

Liu et al. Effect of transcranial magnetic stimulation on treatment effect and immune function. SaudiJ Biol Sci 2022; 29(1): 379-384.

Liu et al. Effects of repetitive transcranial magnetic stimulation combined with music therapy in non-fluent aphasia after stroke: A randomised controlled study. International Journal of Language & Communication Disorders. 2024; 59(3), 1211-1222.

Majdi M, et al. Effect of Repetitive Transcranial Magnetic Stimulation on Meta-Worry and Neuropsychological Functions among Patients with Depression. Iranian Red Crescent Medical Journal 2021; 23(2).

Mansur CG, et al. Placebo effect after prefrontal magnetic stimulation in the treatment of resistant obsessive-compulsive disorder: a randomized controlled trial. International Journal of Neuropsychopharmacology 2011; 14(10): 1389-1397.

Mayer et al. Repetitive Transcranial Magnetic Stimulation as an Add-On Treatment for Cognitive Impairment in Alzheimer's Disease and Its Impact on Self-Rated Quality of Life and Caregiver's Burden. Brain Sciences 2021; 11(6).

Mogg A, Pluck G, Eranti SV, Landau S, Purvis R, Brown RG, et al. A randomized controlled trial with 4-month follow-up of adjunctive repetitive transcranial magnetic stimulation of the left prefrontal cortex for depression. Psychol Med. 2008;38(3):323-33.

Mollica et al. Treatment expectations and clinical outcomes following repetitive transcranial magnetic stimulation for treatment-resistant depression. Brain Stimulation. 2024; 17: 752-759.

Niimi et al. Role of D-serine in the beneficial effects of repetitive transcranial magnetic stimulation in post-stroke patients. Acta Neuropsychiatr. 2020:1-22. (a)

Niimi et al. Effect of repetitive transcranial magnetic stimulation on the kynurenine pathway in stroke patients. Neuroreport. 2020;31(9):629-36. (b)

Ning et al. White matter markers and predictors for subject-specific rTMS response in major depressive disorder. J Affect Disord 2022; 299: 207-214.

Nongpiur et al. Theta-patterned, frequency-modulated priming stimulation enhances low-frequency, right prefrontal cortex repetitive transcranial magnetic stimulation (rTMS) in depression: a randomized, sham-controlled study. J Neuropsychiatry Clin Neurosci. 2011;23(3):348-57.

Norred et al. TMS and CBT-I for comorbid depression and insomnia. Exploring feasibility and tolerability of transcranial magnetic stimulation (TMS) and cognitive behavioral therapy for insomnia (CBT-I) for comorbid major depressive disorder and insomnia during the COVID19 pandemic. Brain Stimul. 2021;14(6):1508-10.

Novák et al. Right ventrolateral and left dorsolateral 10 Hz transcranial magnetic stimulation as an add-on treatment for bipolar I and II depression: a double- blind, randomised, three-arm, sham-controlled study. The World Journal of Biological Psychiatry. 2024; 25(5): 304-316.

Oh & Kim. Adjunctive treatment of bimodal repetitive transcranial magnetic stimulation (rTMS) in pharmacologically non-responsive patients with schizophrenia: a preliminary study. Prog Neuropsychopharmacol Biol Psychiatry. 2011;35(8):1938-43.

Padberg et al. Relation between responses to repetitive transcranial magnetic stimulation and partial sleep deprivation in major depression. J Psychiatr Res. 2002;36(3):131-5.

Pantazatos et al. The timing of transcranial magnetic stimulation relative to the phase of prefrontal alpha EEG modulates downstream target engagement. Brain Stimul. 2023 May-Jun;16(3):830-839.

Picarelli et al. Repetitive transcranial magnetic stimulation is efficacious as an add-on to pharmacological therapy in complex regional pain syndrome (CRPS) type I. J Pain. 2010;11(11):1203-10.

Plewnia et al. Treatment of major depression with bilateral theta burst stimulation: a randomized controlled pilot trial. J Affect Disord. 2014;156:219-23.

Prasser et al. Bilateral prefrontal rTMS and theta burst TMS as an add-on treatment for depression: a randomized placebo controlled trial. World J Biol Psychiatry. 2015;16(1):57-65.

Rapinesi et al. Efficacy of add-on deep transcranial magnetic stimulation in comorbid alcohol dependence and dysthymic disorder: three case reports. Prim Care Companion CNS Disord. 2013;15(1).

Rapinesi et al. Add-on deep Transcranial Magnetic Stimulation (dTMS) for the treatment of chronic migraine: A preliminary study. Neurosci Lett. 2016;623:7-12.
Reddy et al. Brain activation alterations with adjunctive deep transcranial magnetic stimulation in obsessive-compulsive disorder: an fMRI study. CNS Spectr 2022: 1-6.
Rich et al. Repetitive Transcranial Magnetic Stimulation/Behavioral Intervention Clinical Trial: Long-Term Follow-Up of Outcomes in Congenital Hemiparesis. J Child Adolesc Psychopharmacol. 2016;26(7):598-605.
Richter et al. Management of Chronic Tinnitus and Insomnia with Repetitive Transcranial Magnetic Stimulation and Cognitive Behavioral Therapy a Combined Approach. Front Psychol. 2017;8:575.
Rodrigues et al. The Effects of Repetitive Transcranial Magnetic Stimulation on Anxiety in Patients With Moderate to Severe Traumatic Brain Injury: A Post-hoc Analysis of a Randomized Clinical Trial. Frontiers in Neurology 2020; 11.
Rossini et al. Transcranial magnetic stimulation in treatment-resistant depressed patients: a double-blind, placebo-controlled trial. Psychiatry Res. 2005;137(1-2):1-10.
Rotharmel et al. The priming effect of repetitive transcranial magnetic stimulation on clinical response to electroconvulsive therapy in treatment-resistant depression: a randomized, double-blind, sham-controlled study. Psychological Medicine 2022.
Ryan et al. Manipulating Reward Sensitivity Using Reward Circuit-Targeted Transcranial Magnetic Stimulation. Biol Psychiatry Cogn Neurosci Neuroimaging 2022; 7(8): 833-840.
Sampson et al. The use of slow-frequency prefrontal repetitive transcranial magnetic stimulation in refractory neuropathic pain. J ECT. 2011;27(1):33-7.
Sarkhel et al. Adjunctive high-frequency right prefrontal repetitive transcranial magnetic stimulation (rTMS) was not effective in obsessive-compulsive disorder but improved secondary depression. J Anxiety Disord. 2010;24(5):535-9.
Schulze et al. Cognitive safety of dorsomedial prefrontal repetitive transcranial magnetic stimulation in major depression. European Neuropsychopharmacology 2016; 26(7): 1213-1226.
Schutter et al. Increased sensitivity for angry faces in depressive disorder following 2 weeks of 2-Hz repetitive transcranial magnetic stimulation to the right parietal cortex. International Journal of Neuropsychopharmacology 2010; 13(9): 1155-1161.
Senda et al. A Pilot Trial of Stepwise Implementation of Virtual Reality Mindfulness and Accelerated Transcranial Magnetic Stimulation Treatments for Dysphoria in Neuropsychiatric Disorders. Depression and Anxiety. 2023; 9025984.
Seo et al. Adjunctive Low-frequency Repetitive Transcranial Magnetic Stimulation over the Right Dorsolateral Prefrontal Cortex in Patients with Treatment-resistant Obsessive-compulsive Disorder: A Randomized Controlled Trial. Clin Psychopharmacol Neurosci. 2016;14(2):153-60.
Sharma et al. Efficacy of Low-Frequency Repetitive Transcranial Magnetic Stimulation in Ischemic Stroke: A Double-Blind Randomized Controlled Trial. Arch Rehabil Res Clin Transl. 2020;2(1):100039.
Shere et al. Theta burst stimulation in adolescent depression: An open-label evaluation of safety, tolerability, and efficacy. Brain Stimul. 2021;14(4):1051-3.

Singh et al. The safety and efficacy of adjunctive 20-Hz repetitive transcranial magnetic stimulation for treatment of negative symptoms in patients with schizophrenia: A double-blinded, randomized, sham-controlled study. Indian J Psychiatry. 2020;62(1):21-9.

Song et al. High-frequency rTMS over bilateral primary motor cortex improves freezing of gait and emotion regulation in patients with Parkinson's disease: a randomized controlled trial. Frontiers in Aging Neuroscience. 2024; 16:01-09.

Spronk et al. Long-term effects of left frontal rTMS on EEG and ERPs in patients with depression. Clin EEG Neurosci. 2008;39(3):118-24.

Su et al. Add-on rTMS for medication-resistant depression: a randomized, double-blind, sham-controlled trial in Chinese patients. J Clin Psychiatry. 2005;66(7):930-7.

Sureshkumar et al. Role of high-frequency repetitive transcranial magnetic stimulation in augmentation of treatment of bipolar depression. J ect. 2014;30(4):e44-5.

Tadayonnejad et al. Use of right orbitofrontal repetitive transcranial magnetic stimulation (rTMS) augmentation for treatment-refractory obsessive-compulsive disorder with comorbid major depressive disorder. Psychiatry Res 2022; 317: 114856.

Taib et al. Repetitive transcranial magnetic stimulation for functional tremor: A randomized, double-blind, controlled study. Mov Disord. 2019;34(8):1210-9.

Tan et al. Combining transcranial magnetic stimulation and cognitive-behavioral therapy in treatment resistant obsessive-compulsive disorder, Anadolu Psikiyatri Derg. 2015;16(3):180-188.

Tang et al. Analgesic Effects of Repetitive Transcranial Magnetic Stimulation in Patients With Advanced Non-Small-Cell Lung Cancer: A Randomized, Sham-Controlled, Pilot Study. Front Oncol. 2022;12:840855.

Tavares et al. Treatment of Bipolar Depression with Deep TMS: Results from a Double-Blind, Randomized, Parallel Group, Sham-Controlled Clinical Trial. Neuropsychopharmacology. 2017;42(13):2593-601.

Tavares et al. Efficacy, Safety, and Tolerability of Theta-Burst Stimulation in Mixed Depression: Design, Rationale, and Objectives of a Randomized, Double-Blinded, Sham-Controlled Trial. Front Psychiatry. 2020;11:435.

Tavares et al. Treatment of mixed depression with theta-burst stimulation (TBS): results from a double-blind, randomized, sham-controlled clinical trial. Neuropsychopharmacology 2021; 46(13): 2257-2265.

Teti Mayer et al. Repetitive Transcranial Magnetic Stimulation as an Add-On Treatment for Cognitive Impairment in Alzheimer's Disease and Its Impact on Self-Rated Quality of Life and Caregiver's Burden. Brain Sci. 2021;11(6).

Tilbor et al. TMS in combination with a pain directed intervention for the treatment of fibromyalgia - A randomized, double-blind, sham-controlled trial. Journal of Psychiatric Research. 2024; 170:167–173

Tong et al. Impact of Repetitive Transcranial Magnetic Stimulation (rTMS) on Theory of Mind and Executive Function in Major Depressive Disorder and Its Correlation with Brain-Derived Neurotrophic Factor (BDNF): A Randomized, Double-Blind, Sham-Controlled Trial. Brain Sci. 2021;11(6):765.

Torres et al. Retrospective Evaluation of Deep Transcranial Magnetic Stimulation as Add-On Treatment for Parkinson's Disease. Front Neurol. 2015;6:210.

Uygur et al. Successful combination of sleep deprivation therapy and intermittent theta burst stimulation in a patient with recurrent depressive disorder and suicidal ideation. Dusunen Adam-Journal of Psychiatry and Neurological Sciences. 2024; 37(1), 59-60.

Vaithianathan et al. Bilateral sequential theta burst stimulation in depressed veterans with service-related posttraumatic stress disorder: a feasibility study. BMC Psychiatry 2022; 22(1): 81.

Verma et al. Effectiveness of adjunctive repetitive transcranial magnetic stimulation in management of treatment-resistant depression: A retrospective analysis. Indian J Psychiatry. 2018;60(3):329-33.

Vidya et al. Adjuvant Priming Repetitive Transcranial Magnetic Stimulation for Treatment-Resistant Obsessive-Compulsive Disorder: In Search of a New Paradigm! J ECT. 2022;38(1):e1-e8.

Wang et al. Frontoparietal paired associative stimulation versus single-site stimulation for generalized anxiety disorder: a pilot rTMS study. J Psychiatry Neurosci 2022; 47(2): E153-E161.

Wobrock et al. Left prefrontal high-frequency repetitive transcranial magnetic stimulation for the treatment of schizophrenia with predominant negative symptoms: a sham-controlled, randomized multicenter trial. Biol Psychiatry. 2015;77(11):979-88.

Yagci et al. Is the Transcranial Magnetic Stimulation an Adjunctive Treatment in Fibromyalgia Patients? Turkiye Fiziksel Tip Ve Rehabilitasyon Dergisi-Turkish Journal of Physical Medicine and Rehabilitation 2014; 60(3): 206-211.

Yamazaki et al. Laterality of prefrontal hemodynamic response measured by functional near-infrared spectroscopy before and after repetitive transcranial magnetic stimulation: A potential biomarker of clinical outcome. Psychiatry Res 2022; 310: 114444.

Yan et al. [Effect of acupuncture combined with low frequency rTMS on comorbid mild-to-moderate depressive disorder and insomnia: a randomized controlled trial]. Zhongguo Zhen Jiu. 2023; Apr 12;43(4):374-8.

Yu et al. Repetitive transcranial magnetic stimulation promotes response inhibition in patients with major depression during the stop-signal task. Journal of Psychiatric Research 2022; 151: 427-438.

Yuan et al. Effect of Previous Electroconvulsive Therapy on Subsequent Response to Transcranial Magnetic Stimulation for Major Depressive Disorder. Neuromodulation 2020; 23(3): 393-398.

Zangen et al. Repetitive transcranial magnetic stimulation for smoking cessation: a pivotal multicenter double-blind randomized controlled trial. World Psychiatry 2021; 20(3): 397-404.

Zavorotnyy et al. Intermittent theta-burst stimulation moderates interaction between increment of N-Acetyl-Aspartate in anterior cingulate and improvement of unipolar depression. Brain Stimul. 2020;13(4):943-52.

Zendjidjian et al. Resistant bipolar depressive disorder: case analysis of adjunctive transcranial magnetic stimulation efficiency in medical comorbid conditions. Bipolar Disord. 2014;16(2):211-3.

Zhang et al. Add-on rTMS for the acute treatment of depressive symptoms is probably more effective in adolescents than in adults: Evidence from real-world clinical practice. Brain Stimul. 2019;12(1):103-9.

Zhang et al. Comparative efficacy of add-on rTMS in treating the somatic and psychic anxiety symptoms of depression comorbid with anxiety in adolescents, adults, and elderly patients-A real-world clinical application. J Affect Disord. 2020;276:305-11.
Zhang et al. An Open-label Trial of Adjuvant High-frequency Left Prefrontal Repetitive Transcranial Magnetic Stimulation for Treating Suicidal Ideation in Adolescents and Adults With Depression. J ECT. 2021;37(2):140-6. (a)
Zhang et al. Clinical observation of acupuncture plus repetitive transcranial magnetic stimulation in the treatment of post-stroke insomnia, J J. Acupunct. Tuina Sci. 2020;18(2):122-128. (b)
Zhang et al. Task-related functional magnetic resonance imaging-based neuronavigation for the treatment of depression by individualized repetitive transcranial magnetic stimulation of the visual cortex. Science China-Life Sciences 2021; 64(1): 96-106.

***Is protocol, conference paper, book chapter without new data, secondary analysis, or abstract***

Abou El-Magd et al. Repetitive Transcranial Magnetic Stimulation With and Without Internet-Delivered Cognitive-Behavioral Therapy for the Treatment of Resistant Depression: Protocol for Patient-Centered Randomized Controlled Pilot Trial. JMIR Res Protoc. 2020;9(10):e18843.
Adu et al. Repetitive Transcranial Magnetic Stimulation with and without Internet-Delivered Cognitive Behavior Therapy for the Treatment of Resistant Depression: Patient-centered Randomized Controlled Pilot Trial. European Psychiatry. 2023; 66, S416-S416.
Bulteau et al. Cost utility analysis of curative and maintenance repetitive transcranial magnetic stimulation (rTMS) for treatment-resistant unipolar depression: a randomized controlled trial protocol. Trials. 2020;21(1):312.
Canali et al. Depression, cortical excitability and sleep deprivation: a TMS/EEG study. European Neuropsychopharmacology 2012; 22:S276-S277.
Dalhuisen et al. rTMS combined with CBT as a next step in antidepressant non-responders: a study protocol for a randomized comparison with current antidepressant treatment approaches. BMC Psychiatry. 2022; 22(1):88.
Davis et al. Network-level dynamics underlying a combined rTMS and psychotherapy treatment for major depressive disorder: An exploratory network analysis. Int J Clin Health Psychol. 2023;23(4):100382.
Demitrack et al. Transcranial Magnetic Stimulation (TMS) in the Treatment of Pharmacoresistant Major Depression: Examination of Cognitive Function During Acute Treatment, Biol Psychiatry. 2009;65(8):221S-222S.
Ekpo et al. Concurrent fMRI-Guided rTMS and Cognitive Therapy for the Treatment of Major Depressive Episodes. Journal of Ect. 2023; 39(3): E18-E19.
Haque & Malik. Efficacy of Repetitive Transcranial Magnetic Stimulation With Concurrent Cognitive Behavioral Therapy on Treatment-Resistant Depression. J ECT. 2017;33(3):212-212.
He et al. Treatment of depression using sleep electroencephalogram modulated repetitive transcranial magnetic stimulation. Eur Psychiatry. 2011;26.
Hill et al. Characterizing TMS-Related Oscillatory Dynamics in Treatment Resistant Depression Before and After Convulsive Therapy: A TMS-EEG Study. Biological Psychiatry 2021; 89(9): S193-S193.

Hu & Lisanby. Cognitive Behavioral Strategies During Transcranial Magnetic Stimulation in Obsessive-Compulsive Disorder and Concomitant Major Depressive Disorder, J ECT. 2015;31(3):E35-E36.

Iseger et al. Optimizing TMS Treatment for Depression Using Cardiac Response With Neuro-Cardiac-Guided-TMS (NCG TMS), Biol Psychiatry. 2018;83(9):S55-S55.

Jha et al. Long Term Efficacy Of Brain Spect Assisted Repetitive Transcranial Magnetic Stimulation (RTMS) In Treatment Resistant Major Depressive Disorder- One Year Follow-up Study, Indian J Psychiatry. 2017; 59(6):S188-S189.\

Jin et al. EEG Synchronized TMS For Individualized Therapy in Major Depressive Disorder, Eur Psychiatry. 2011;26.

Kujovic et al. Theta burst stimulation add on to dialectical behavioral therapy in borderline-personality-disorder: methods and design of a randomized, single-blind, placebo-controlled pilot trial. European Archives of Psychiatry and Clinical Neuroscience. 2024; 274(1): p. 87-96.

Lee et al. A REPETITIVE TRANSCRANIAL MAGNETIC STIMULATION (rTMS) SERVICE FOR THE TREATMENT OF DEPRESSIVE DISORDERS. Australian and New Zealand Journal of Psychiatry 2014; 48: 59-59.

Lee et al. Combination Theta-Burst Stimulation and Cognitive Training for Youth Depression, Biol Psychiatry. 2019;85(10):S181-S181.

Lee et al. Modified trains of rTMS pulses, that take account of background EEG, can be used in developing stimulus parameters for the treatment of depression, Int. J. Neuropsychopharmacol. 2006;9:S179-S179.

Lee et al. Repetitive transcranial magnetic stimulation with EEG-dependent stimulus timing in the treatment of depression, Eur Neuropsychopharmacol. 2009;19:S478-S478.

Leuchter & Hunter. A resting-state quantitative electroencephalographic (qEEG) biomarker of treatment outcome during repetitive transcranial magnetic stimulation (rTMS) treatment of major depressive disorder (MDD). Bipolar Disorders 2016; 18: 100-100.

Leuchter et al. Effectiveness of Synchronized Transcranial Magnetic Stimulation (sTMS) for Treatment of Major Depression. Biological Psychiatry 2014; 75(9): 120S-121S.

Maslenikov et al. Repetitive transcranial magnetic stimulation (rTMS) versus antidepressants as an add-on treatment for depression in schizophrenia. European Neuropsychopharmacology 2019; 29: S230-S231.

Oathes et al. Effects of Stimulation Site, Context, and Trauma History on Response to rTMS Treatment Among Patients With PTSD or Depression With Trauma. Biol Psychiatry. 2022;91(9):S36-S36.

Stikhina et al. Transcranial magnetic stimulation in neurotic depression. Zhurnal Nevropatologii I Psikhiatrii Imeni S S Korsakova.1999; 99(10): 26-29.

Tang et al. Repetitive transcranial magnetic stimulation for depression after basal ganglia ischaemic stroke: protocol for a multicentre randomised double-blind placebo-controlled trial. BMJ Open. 2018;8(2):e018011.

Tynan et al. Neuromodulation-Assisted Psychotherapy: Can Transcranial Magnetic Stimulation Increase The Efficacy Of Acceptance And Commitment Therapy For Comorbid Chronic Pain And Depression? The Journal of Pain. 2023; 24(4):63

Uribe et al. The Effects of Repetitive Transcranial Magnetic Stimulation (rTMS) on Rumination in Major Depressive Disorder (MDD), Biol Psychiatry. 2020;87(9):S449-S450.

Vaishnavi & Brammer. Transcranial Magnetic Stimulation of the Left Dorsolateral Prefrontal Cortex and Response to Negative Faces in Major Depression, Journal of Neuropsychiatry and Clinical Neurosciences. 2022;34(3):300-300.
Walpoth et al. Repetitive transcranial magnetic stimulation (rTMS) as an add-on therapy in depression: A prospective, monocenter, randomized, double-blind, sham-controlled trial. European Neuropsychopharmacology 2003; 13: S176-S177.
Wen et al. Theta-Burst Stimulation Combined With Virtual-Reality Reconsolidation Intervention for Methamphetamine Use Disorder: Study Protocol for a Randomized-Controlled Trial, Frontiers in Psychiatry. 2022;13.

***Depressive symptom severity not assessed***

Carmi et al. Clinical and electrophysiological outcomes of deep TMS over the medial prefrontal and anterior cingulate cortices in OCD patients. Brain Stimul. 2018;11(1):158-65.
Chen et al. The role of dorsolateral prefrontal cortex on voluntary forgetting of negative social feedback in depressed patients: A TMS study, Acta Psychologica Sinica. 2021;53(10);1094-1104.
Rabey et al. Repetitive transcranial magnetic stimulation combined with cognitive training is a safe and effective modality for the treatment of Alzheimer's disease: a randomized, double-blind study. Journal of Neural Transmission. 2013;120(5):813-9.
Ross et al. Simultaneous aerobic exercise and rTMS: Feasibility of combining therapeutic modalities to treat depression. Brain Stimul. 2018;11(1):245-6.
Sokhadze et al. Neuromodulation integrating rTMS and neurofeedback for the treatment of autism spectrum disorder: an exploratory study. Appl Psychophysiol Biofeedback. 2014;39(3-4):237-57.

***Specialized NIBS protocol (e.g., bilateral protocols or EEG-guided), but not combined with psychological task or intervention***

Kavakbasi et al. Vagus Nerve Stimulation Combined With Alternating Synchronized and Nonsynchronized Intermittent Theta Burst Stimulation in Difficult-to-Treat Depression. J ect. 2024; 40(1): p. 62-63.
Leuchter et al. Efficacy and Safety of Low-field Synchronized Transcranial Magnetic Stimulation (sTMS) for Treatment of Major Depression. Brain Stimul. 2015 Jul-Aug;8(4):787-94.
McDonald et al. Combination rapid transcranial magnetic stimulation in treatment refractory depression. Neuropsychiatr Dis Treat. 2006;2(1):85-94.
Li et al. Preliminary Research on Depression Treatment: Combination of Transcranial Magnetic Stimulation and MRI-Guided Low-Intensity Focused Ultrasound Pulsation. Journal of Medical Imaging and Health Informatics 2020; 10(3): 677-680.
Price et al. The use of background EEG activity to determine stimulus timing as a means of improving rTMS efficacy in the treatment of depression: a controlled comparison with standard techniques. Brain Stimul. 2010;3(3):140-52.
Robertson & Mortimer. Quantitative EEG (qEEG) guided transcranial magnetic stimulation (TMS) treatment for depression and anxiety disorders: An open, observational cohort study of 210 patients. J Affect Disord. 2022;308:322-327.

Sreepada et al. Dual stimulation with tDCS-iTBS as add-on treatment in recurrent depressive disorder - a case report. Brain Stimulation 2020; 13(3): 625-626.
Zrenner et al. Brain oscillation-synchronized stimulation of the left dorsolateral prefrontal cortex in depression using real-time EEG-triggered TMS, Brain Stimul. 2020;13(1):197-20.

##### *rTMS was not applied*

Canali et al. Changes of cortical excitability as markers of antidepressant response in bipolar depression: preliminary data obtained by combining transcranial magnetic stimulation (TMS) and electroencephalography (EEG). Bipolar Disorders. 2014;16(8): 809-819.
Kaneko et al. Neuroplasticity of the left dorsolateral prefrontal cortex in patients with treatment-resistant depression as indexed with paired associative stimulation: a TMS-EEG study. Cereb Cortex. 2024; 34(2).
Lissemore et al. An inverse relationship between cortical plasticity and cognitive inhibition in late-life depression. Neuropsychopharmacology 2019; 44(9): 1659-1666.
Ross et al. Acute effects of aerobic exercise on corticomotor plasticity in individuals with and without depression. Journal of Psychiatric Research. 2024; 176, 108-118.
Salehinejad et al. Sleep-dependent upscaled excitability, saturated neuroplasticity, and modulated cognition in the human brain. Elife. 2022;11:e69308.

##### *Healthy participants recruited only*

Dalhuisen et al. Studying additive effects of combining rTMS with cognitive control training: a pilot investigation. Frontiers in Human Neuroscience. 2023;17; 1201344.
Grosshagauer et al. Chronometric TMS-fMRI of personalized left dorsolateral prefrontal target reveals state-dependency of subgenual anterior cingulate cortex effects. Mol Psychiatry. 2024.

##### *Could not retrieve full text*

Yin et al. Acupuncture combined with repetitive transcranial magnetic stimulation for post-stroke depression: a randomized controlled trial. Zhongguo Zhen Jiu 2022; 42(11): 1216-20.

#### Supplementary Text

##### Supplementary Text 1. *Effect sizes based on change scores of within-group and between-group comparisons*

Most included studies measured depression severity with standardized scales to assess the effects of active or sham combinations of rTMS with psychological method (e.g., psychotherapy, bright light therapy, psychophysical tasks), and reported the baseline and immediate endpoint means, standard deviations, and sample sizes.

To compute effect sizes, we used custom scripts written in Python (version 3.11.5) with NumPy and panda libraries in a Jupyter Notebook environment. We computed standardized mean differences of change scores for individual study comparisons:

$$d = \frac{\bar{x}_{active\ change} - \bar{x}_{control\ change}}{SD_{pooled\ change}} \quad \text{[Equation 1]}$$

Where $d$ is standardized mean difference (Cohen's d), $\bar{x}_{active\ change}$ is the average change score for the active group and $\bar{x}_{control\ change}$ is the average change score for the control group (or for within-group comparisons, the baseline average change scores is subtracted from the endpoint average change scores), and $SD_{pooled\ change}$ is the pooled standard deviation of the change scores from both groups (or just one group for within-group comparisons) (Higgins et al., 2019). We then converted this estimate to Hedges' g to correct for potential bias due to small sample sizes (Hedges et al., 1981):

$$g = d * J = d * (1 - \frac{3}{(4*df)-9}) \quad \text{[Equation 2]}$$

Where $J$ is the bias correction factor, and $df$ is the degrees of freedom, which equals n-1 for within-group comparisons and $n_1+n_2$-2 for between group comparisons (Hedges et al., 1981).

*Controlled Effect Sizes*

This extracted data was used to compute the standardize mean differences (SMDs) of change scores for two groups being compared (e.g., active combination treatments – sham combination treatments). Below is pseudocode of the equations ran in custom python scripts; used after importing *numpy* and *pandas* libraries.

```
Function name: "calculate_controlledEffectSizes"
    Required input:
        active_premeans, active_preSD, active_postmeans, active_postSD, active_n
        sham_premeans, sham_preSD, sham_postmeans, sham_postSD, sham_n
        prepost_r

    Mean score changes:
        ChangeScore_active = active_postmeans - active_premeans
        ChangeScore_sham = sham_postmeans - sham_premeans

    Difference in mean score changes:
        ChangeScore_diff = ChangeScore_active - ChangeScore_sham

    Standard deviations of the above mean score changes:
        SD_ChangeScore_active = sqrt(active_preSD^2 + active_postSD^2 - 2 * prepost_r * active_preSD * active_postSD)
        SD_ChangeScore_active = sqrt(sham_preSD^2 + sham_postSD^2 - 2 * prepost_r * sham_preSD * sham_postSD)

    Pooled standard deviation of the changes:
        pooled_SD = sqrt(((active_n - 1) * SDChange_active^2 + (sham_n - 1) * SDChange_sham^2) / (active_n + sham_n - 2))

    Standardized difference in means (Cohen's d):
        SMD = ChangeScore_diff / pooled_SD

    Standard error of the standardized difference:
        SMD_SE = sqrt(1 / active_n + 1 / sham_n + SMD ^2 / (2 * (active_n + sham_n)))

    Return effect size (SMD = Cohen's d) and its standard error.
```

These SMDs were then converted to Hedges' g with the correction factor:

```
Function name: "SMD_to_g"
    Required input:
        SMD, SMD_SE, active_n, sham_n

    Degrees of freedom and correction factor (J):
        df = active_n + sham_n - 2
        J = 1 - (3 / (4 * df - 1))

    Hedges' g and its standard error:
        g = SMD * J
        g_SE = SMD_SE * J

    Return corrected effect size (Hedges' g) and its standard error.
```

*Uncontrolled Effect Sizes*

Effect sizes of active combinations of rTMS with psychological methods were also pooled.

```
Function name: “calculate_wg_hedgesg” # “wg” dis-abbreviates to ‘within group’
  Required input:
    baseline_mean, baseline_sd, endpoint_mean, endpoint_sd, prepost_r, n

  Within-group mean differences
    wg_diff = endpoint_mean - baseline_mean
    wg_diff_sd = sqrt(baseline_sd ^2 + endpoint_sd ^2 - 2 * prepost_r * baseline_sd * endpoint_sd)
    wg_diff_se = wg_diff_sd / sqrt(n)

  Standardized mean difference (SMD)
    wg_SMD = wg_diff / (wg_diff_sd / sqrt(2 * (1 - prepost_r)))
    wg_SMD_se = sqrt((1 / n) + (wg_SMD^2 / (2 * n))) * sqrt(2 * (1 - prepost_r))

  Corrected SMD (g)
    df = n - 1
    j = 1 - (3 / (4 * df - 1))
    wg_g = wg_SMD * j
    wg_g_SE = wg_SMD_se * j

  Return corrected effect size (Hedges' g) and its standard error.
```

*Special Cases*

Few included studies did not provide means or standard deviations, or reported SMDs. These were handled case-by-case.

Two included studies (Osuch et al., 2009, reference number 49; Ross et al., 2023, reference number 50) provided the within-group SMD (Cohen's d) of their findings. The correction factor was applied to SMD to estimate Hedges' g.

```
Corrected SMD (g)
    df = n - 1
    j = 1 - (3 / (4 * df - 1))
    g = SMD * j
    g_SE = SMD_se * j

Return corrected effect size (Hedges' g) and its standard error.
```

One included study (Fryml et al., 2019, reference number 42) reported the interaction F-statistic of time x treatment. We used the sample sizes for each group and the reported F-statistic to compute the SMD and its standard error, which was subsequently converted to Hedges' g, as above.

```
SMD = sqrt(F * ((n1 + n2) / (n1 * n2)))
SMD_se = sqrt((1 / n1) + (1 / n2) + (SMD^2 / (2 * (n1 + n2))))
```

**Supplementary Text 2.** ***Meta-analysis results of depression change scores across disorders***

Four studies compared groups receiving [active rTMS + active PSYC] versus [active rTMS + sham PSYC]. The figures below show the forest and funnel plots for this analysis and discussed in the main text.

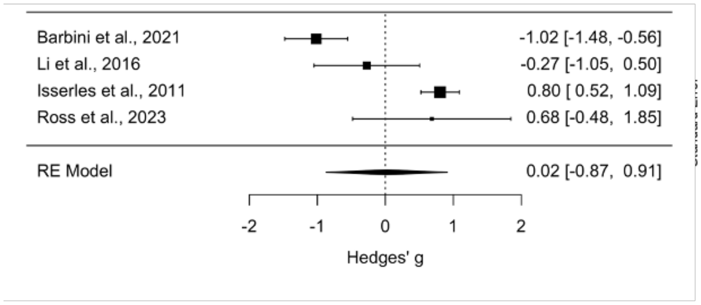


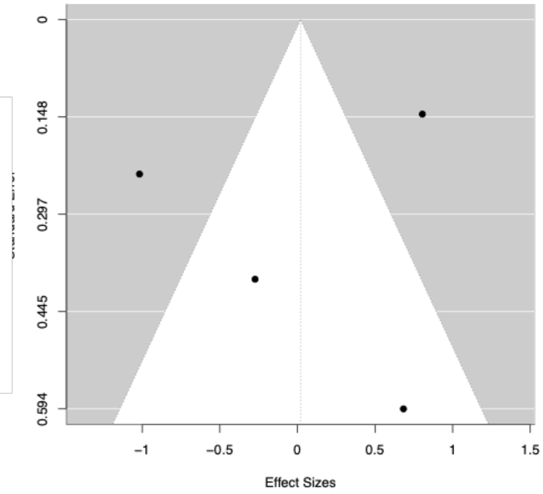

**Supplementary Text 3:** ***Secondary meta-analysis: left DLPFC treatment for depressive disorders***

Within-group comparisons of [active rTMS + active PSYC] at endpoint minus baseline change scores included 14 studies (16 datasets) where the left DLPFC of patients with depressive conditions was targeted. The figures below show the forest and funnel plots for this analysis and discussed in the main text.

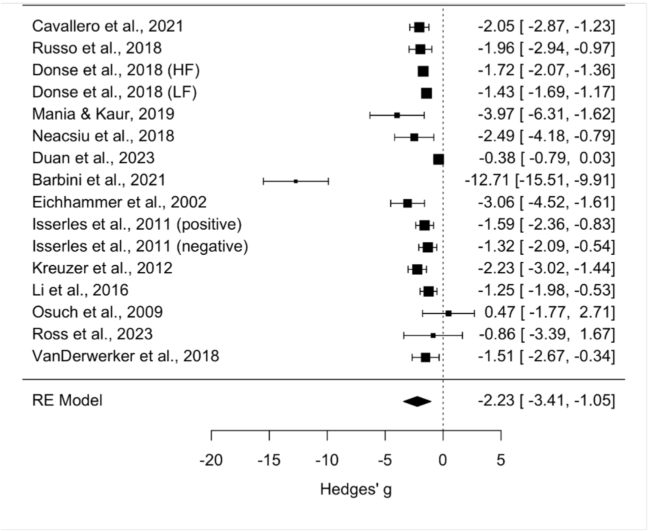


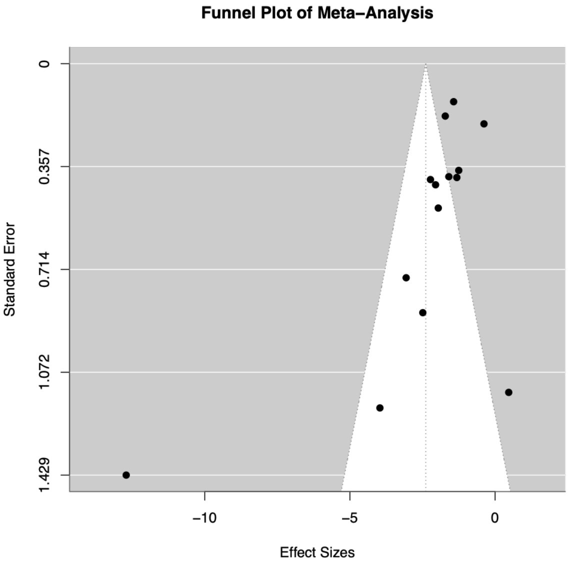


Five studies compared [active rTMS + active PSYC] versus [sham rTMS + active PSYC] and targeted the left DLPFC of patients with depressive conditions.

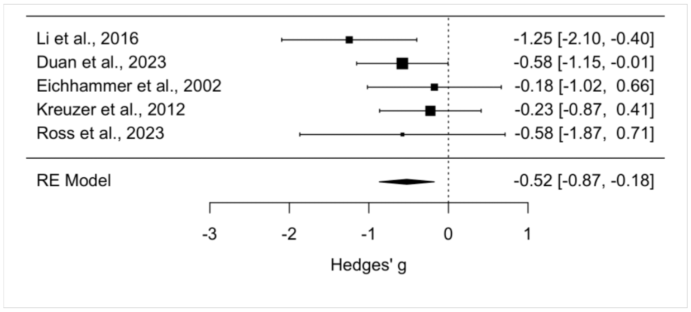


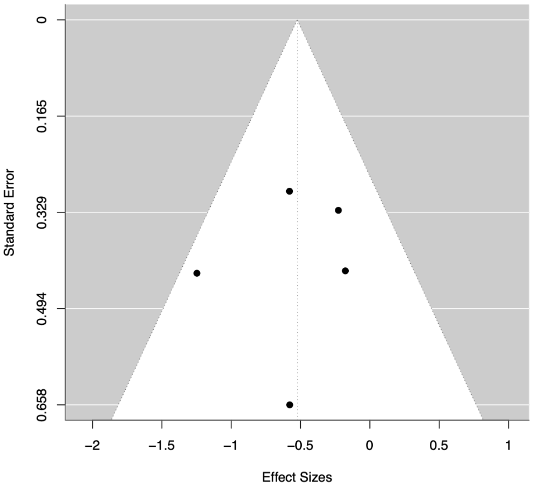

**Supplementary Text 4:** ***Power analysis methods and results***

To estimate the sample size needed to obtain sufficient power ($\beta$=0.80, given $\alpha = 0.05$) to investigate the hypothesis that combining rTMS with PSYC is a superior treatment (compared to sham conditions), we used the results of representative studies in simulation-based power analysis. The specific results were the average change scores per group. The simulated analysis was basic and reflects published work: a one-way ANOVA with four groups (both active treatments group, active rTMS with sham PSYC, sham rTMS with active PSYC, and both sham treatments group); the dependent variable was change scores randomly sampled from a normal distribution with mean and standard deviation imputed from efficacy data reported in included studies. If the ANOVA was significant, Tukey's Honest Significant Difference (HSD) post hoc test was conducted to compare pairwise differences among treatment groups, while controlling for multiple comparisons within a simulation. The custom Python script counted how the frequency that "both active" group was significantly better than the other groups, reflecting the definition of power and investigating the research hypothesis that active combinations were significantly better than all other treatments.

Simulations assumed a set of sample sizes ranging from 4 (one patient per group) to 240 (60 patients per group), with 10,000 iterations for each sample size. This analysis assumes a normal distribution of change scores and independence among observations. Power was formulated as follows, representing the probability of correctly rejecting the null hypothesis when it is false:

$$power = \frac{frequency \text{ "both\_active" } was\ best}{total\ number\ of\ simulations} \qquad \text{[Equation 3]}$$

Power analysis for testing the hypothesis that [active rTMS + active PSYC] was significantly better than [sham rTMS + active PSYC] and [sham rTMS + sham PSYC], but not compared to [active rTMS + sham PSYC]. The figure below also shows the threshold to achieve 80% power; the table below the figure shows values corresponding to specific sample sizes. Alpha is 0.05.

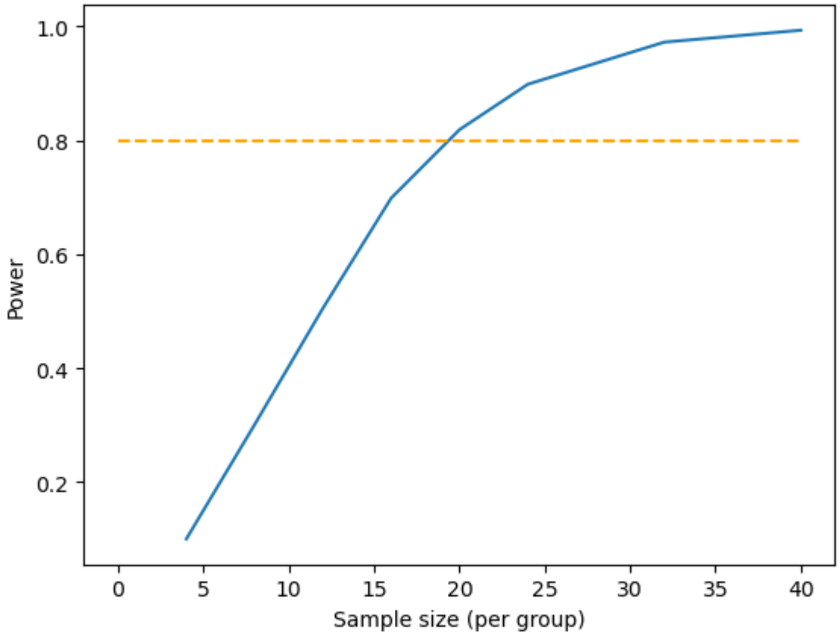


| Sample Size (per group) | Power (%) |
|---|---|
| 4 | 10.05 |
| 8 | 30.12 |
| 12 | 50.54 |
| 16 | 69.87 |
| 20 | 81.88 |
| 24 | 89.85 |
| 32 | 97.25 |
| 40 | 99.34 |

Power analysis for testing the hypothesis that the both active treatment is better than all active and sham comparators (including [active rTMS + sham PSYC]). The figure below also shows the threshold to achieve 80% power; the table below shows values corresponding to specific sample sizes. Alpha is 0.05.

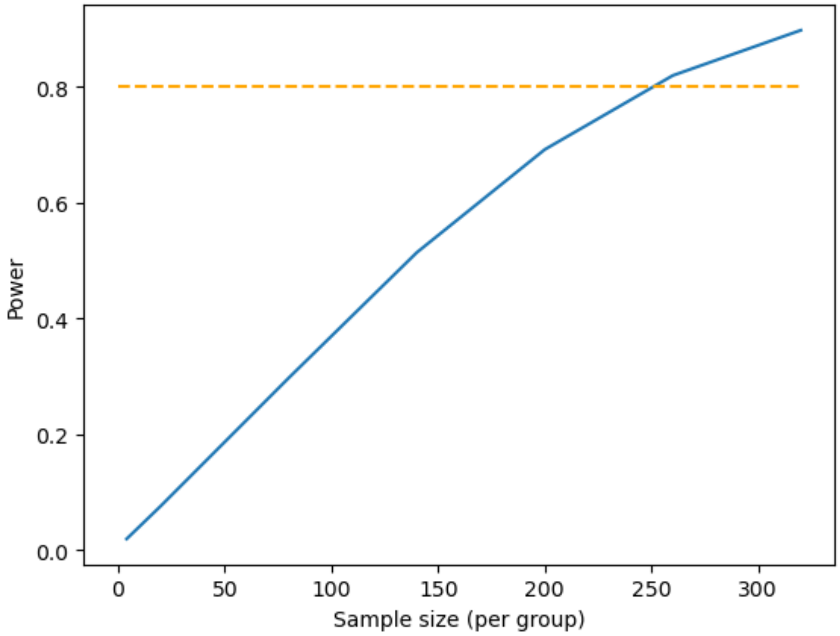


| Sample Size (per group) | Power (%) |
|---|---|
| 4 | 1.95 |
| 20 | 7.63 |
| 80 | 29.74 |
| 140 | 51.38 |
| 200 | 69.16 |
| 260 | 81.93 |
| 320 | 89.74 |
